# A foundation model of wearable pulse oximetry reveals physiological signatures of health and cardiometabolic risk

**DOI:** 10.64898/2026.07.01.26356992

**Authors:** Sarah Kohn, Guy Lutsker, Alon Diament, Smadar Shilo, Adam Gabet, Gil Sasson, Gili Wolf, Adva Wolf, Anastasia Godneva, Adina Weinberger, Hagai Rossman, Eran Segal

## Abstract

While Photoplethysmography (PPG) is established as a noninvasive optical tool for monitoring heart rate and oxygen saturation, its high-resolution pulsatile blood volume waveforms contain rich physiological data that extend far beyond conventional vital signs.

We introduce PulseOx-FM, a foundation model, trained using self-supervised learning on 6,995,558 segments of pulse oximetry signals collected during 42,282 overnight sleep monitoring recordings of 10,704 adult participants in the Human Phenotype Project (HPP).

Using chronological age as a global health benchmark, PulseOx-FM significantly outperformed existing open-source and proprietary feature extraction methods while demonstrating robust generalization in an external out-of-distribution cohort. PulseOx-FM representations predicted 71 phenotypic targets spanning cardiometabolic, and neuropsychiatric domains beyond age, sex and body mass index. Applied in the same way to 3,721 intensive-care patients, two-minute windows of routine bedside pulse oximetry from the first day of admission improved the prediction of survival beyond age, sex and the heart rate and oxygen saturation the same device reports. Nightly embeddings further tracked next-day glycemic, dietary and activity-based state within individuals, dissociating this signal from sleep architecture alone. This next-day glycemic signal was predominantly a direct physiological effect, not explained by next-day dietary intake.

These findings suggest that PulseOx-FM provides a generalizable framework for encoding physiological patterns from sleep, offering a non-invasive tool for global health risk stratification and precision medicine.

## Introduction

Photoplesthysmography (PPG) is a noninvasive, optical sensing technology that measures pulsatile blood volume variations in the peripheral vasculature by emitting light into tissue and detecting changes in light absorption ^1^. Embedded in consumer wrist devices, fingertip monitors, and hospital oximeters alike, it is the most scalable continuous physiological sensor ^2^. In clinical medicine, it serves as the core mechanism powering pulse oximetry signals for continuous heart rate and blood oxygen saturation (SpO_2_) monitoring ^3^, yet the PPG waveform rich dynamics encoding cardiovascular, autonomic, and respiratory state remains largely uninterpreted ^4^. Unlocking this potential requires both a learning framework and a training context offering physiological variation, reproducibility, and availability at scale ^2^. These are conditions that sleep satisfies. As a nightly period of relative behavioral quiescence, sleep allows the body’s regulatory systems to operate in a consistent, unconfounded manner (with fewer movement artifacts and less behavioral variability than daytime recordings). During this period, PPG can be captured at scale, leveraging its widespread use across clinical diagnostics, research cohorts, and consumer wearables ^3^.

Standard deep learning applications for continuous physiological time series, and for PPG specifically, remain highly constrained by task-specific supervision, such as single-channel sleep staging frameworks ^5^. The emergence of self-supervised learning (SSL) has enabled the development of foundation models that extract meaningful representations from large, unlabeled datasets ^6^. Recent self-supervised work has shown success in retinal imaging ^7^, pathology ^8^, and continuous glucose monitoring ^9^, and has even begun to reframe PPG and related wearable biosignals as general-purpose physiological encoders rather than task-specific vital-sign streams. Large-scale efforts, including self-supervised PPG and ECG encoders trained on approximately 141,000 participants in the Apple Heart and Movement Study ^10^, and open PPG foundation models such as PaPaGei ^11^, Pulse-PPG ^12^, and GPT-PPG ^13^, together with ECG-guided models such as AnyPPG ^14^, and deployment work such as PPG-Distill ^15^, collectively demonstrate rapid progress in PPG representation learning. In parallel, PPG-derived aging clocks such as PpgAge ^16^ and AI-PPG ^17^ establish that wrist PPG encodes chronological and cardiovascular age, while multimodal sleep foundation models such as SleepFM ^18^ illustrate the breadth of disease prediction achievable from a single overnight recording. Although comprehensive, SleepFM relies on full polysomnography (PSG), and the requirement for brain activity (EEG), heart (ECG), and muscle tone (EMG) monitoring makes these PSG-based models difficult to scale for long-term population health monitoring. PPG-based approaches overcome this limitation, requiring only a consumer wearable and no specialist setup, and thus surpass even home-based sleep studies in accessibility. This creates a significant technological divide: existing PPG models are highly scalable but limited to niche diagnostic tasks, while comprehensive sleep models are broadly predictive but clinically cumbersome. PulseOx-FM addresses this division: it learns from overnight pulse-oximetry signals alone, in a deeply phenotyped longitudinal cohort, linking a scalable optical signal to vascular aging, cardiometabolic state, incident hypertension, and within-person next-day metabolic variation, with external transfer to intraoperative and intensive-care recordings.

Recent phenome-wide association studies (PheWAS) using predefined sleep biomarkers, from homebased sleep apnea tests mainly focusing on PPG / Peripheral Arterial Tonometry (PAT) monitoring, have revealed extensive links between nocturnal physiology and several body systems, particularly in predicting glycemic parameters and blood lipids ^19^. However, these biomarker extraction methods rely on a finite set of derived characteristics, that prioritize specific diagnostic outputs such as the apnea-hypopnea index (AHI), and may therefore overlook richer physiological information encoded in the raw waveform dynamics. Hand-crafted features are inherently domain-specific, engineered for a particular diagnostic target ^20,21^, they tend to be implicitly overfitted to that domain, which is reflected empirically in their difficulty to generalize beyond the tasks they were designed for.

Here we introduce PulseOx-FM, a transformer-based foundation model pretrained on large pulse oximetry data from the Human Phenotype Project (HPP) to extract broad health signatures from raw peripheral pulse-oximetry signals. The study was structured chronologically to transition from model development to clinical application (Fig. 1). By employing a masked auto-encoder (MAE) objective, PulseOx-FM learns latent representations of physiology without the need for manual labels. We evaluate its performance across diverse tasks, from signal reconstruction (Extended Data Fig. 1, Supp. Fig. 1) to the prediction of chronic disease including cardiovascular, mental and metabolic outcomes. Notably, evaluation on an external out-of-domain dataset reveals that a model pretrained purely on ambulatory, sleep PPG data successfully captures fundamental physiological features that generalize to the acute, intraoperative state under general anesthesia. This capacity to translate across vastly different clinical contexts provides compelling evidence that PulseOx-FM learns intrinsic, systemic representations of human physiology rather than just dataset-specific artifacts, highlighting its potential as a scalable, universally generalizable tool for continuous health monitoring and personalized risk assessment.

**Fig. 1.**
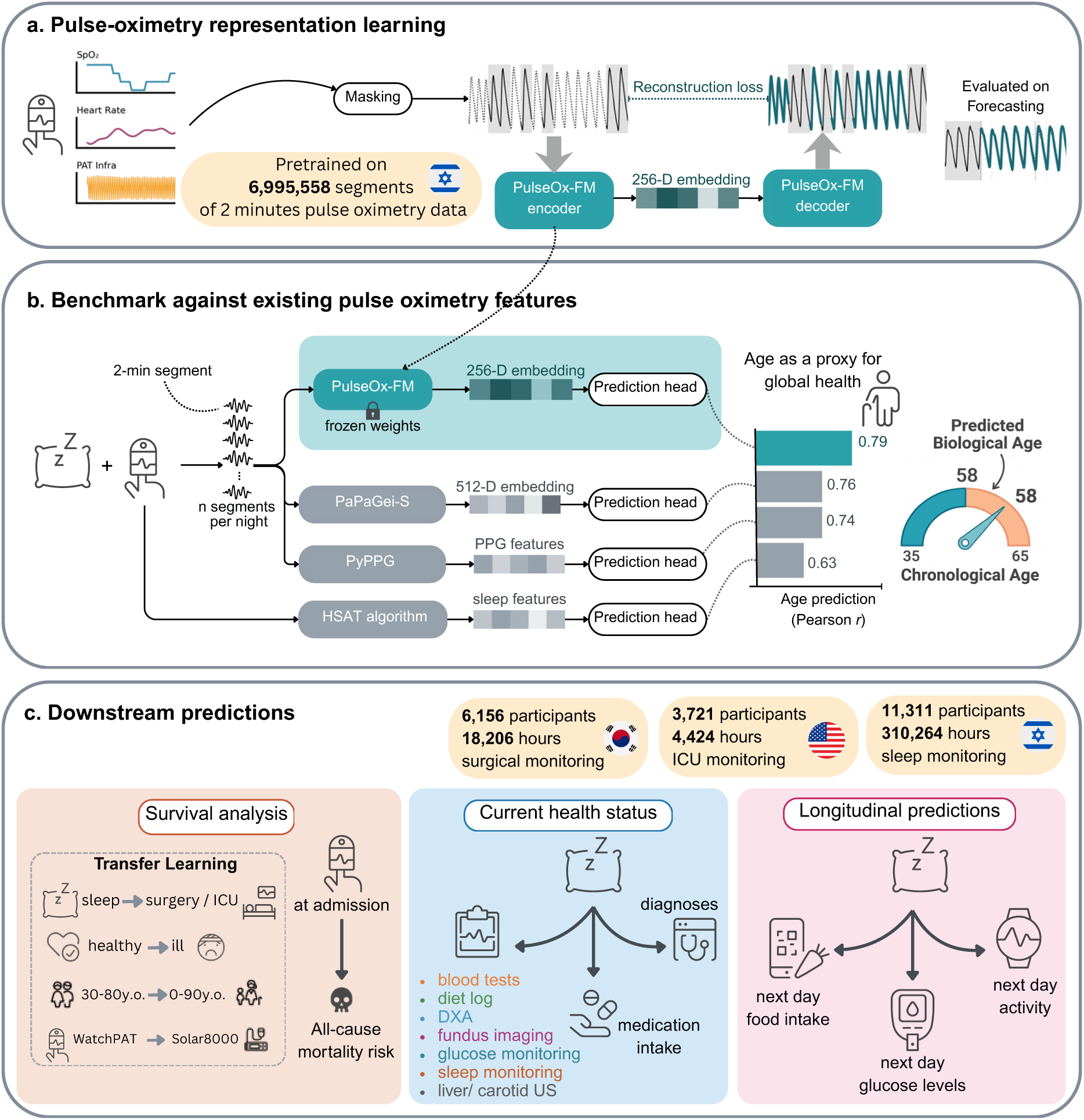
Overview of the PulseOx-FM framework and study design. **a**, Schematic of the PulseOx-FM architecture, which uses a transformer-based masked auto-encoding objective to learn 256-dimensional latent representations from 2-minute segments of raw pulse oximetry data: oxygen saturation (SpO_2_), heart rate and the peripheral arterial tonometry (PAT) infrared waveform. The pretraining corpus contains 6,995,558 two-minute segments from the Human Phenotype Project (HPP) sleep-monitoring cohort. **b**, Benchmarking of PulseOx-FM against a peer foundation model (PaPaGei-S), photoplethysmography (PPG) features (PyPPG) and the features of the home sleep apnoea test (HSAT) algorithm. Frozen embeddings, pooled across the *n* two-minute segments of each night, are passed to a lightweight prediction head. Chronological age, used as a proxy for global health status, is predicted more accurately from PulseOx-FM embeddings than from any comparator. **c**, Downstream tasks in three independent cohorts, HPP (sleep monitoring, Israel), VitalDB (surgical monitoring, South Korea) and MIMIC-III (intensive-care unit (ICU) monitoring, USA). The tasks are survival analysis, in which transfer learning from sleep to surgery and ICU, from healthy to ill, and across devices is used to predict all-cause mortality risk from monitoring early in the admission; current health status (blood tests, diet, dual-energy X-ray absorptiometry (DXA), fundus imaging, glucose and sleep monitoring, liver and carotid ultrasound (US), medication intake and diagnoses); and longitudinal predictions (next-day food intake, glucose levels and activity).

The aim of this study is to test whether a model trained without labels on routine overnight pulse oximetry learns a general-purpose representation of an individual’s health. We address three subaims: first, to develop the model and benchmark it against existing feature-based approaches and against PaPaGei-S, an open PPG foundation model, using chronological age as a global readout, in the HPP and in external surgical and intensive-care cohorts; second, to test whether its embeddings carry information on current health beyond age, sex and body mass index; and third, to test whether they carry forward-looking information, namely two-year incident hypertension and next-day glycaemic state. Finally, we evaluate one clinical use case, early mortality risk stratification from bedside pulse oximetry recorded during the first day of an intensive-care admission.

## Results

### Study design and cohorts characteristics

PulseOx-FM was pretrained on physiological monitoring data from the HPP, a large-scale longitudinal study ^22^. The dataset was collected using the FDA-cleared WatchPAT home sleep diagnostics device (ZOLL Itamar), which noninvasively tracks peripheral dynamics alongside systemic vitals. The model leverages three of the device’s synchronous channels: SpO_2_, pulse rate and the peripheral pulsatile blood volume waveform. This waveform is captured via the device’s proprietary Peripheral Arterial Tonometry (PAT) finger sensor; while PAT explicitly measures autonomic-driven changes in arterial tone, it shares the same underlying optical sensing principles as standard PPG, yielding a highly analogous pulsatile blood volume signal. In total, PulseOx-FM was trained, validated, and tested on 9,854,117 2-minute segments across two distinct clinical settings. The primary development cohort from the HPP study included 11,311 participants, providing 45,068 overnight recordings, amounting to 310,264 hours of sleep data. To evaluate the model’s robustness and generalizability, we performed external validation using the VitalDB cohort ^23^, which comprises intraoperative monitoring data from 6,156 participants collected during surgery, totaling 18,206 hours. A third, intensive-care cohort was drawn from the MIMIC-III Waveform Database Matched Subset ^24^, which links bedside-monitor recordings of adult intensive-care patients in Boston, USA, to their clinical records. To ensure that no input was recorded later in the stay or during a later admission, we restricted it to recordings made within the first day of the first admission (Methods), giving 3,721 patients and 4,424 hours of monitoring. Together, the three cohorts comprise 21,188 participants from three countries, recorded in three acquisition contexts (home sleep monitoring, general anaesthesia and critical care) on three classes of device. Comprehensive demographic information and recruitment protocols for all cohorts are summarized in Table 1 and the Methods section, and the medication and diagnosis categories analysed are listed in Supp. Tables 1 and 2.

**Table 1.**
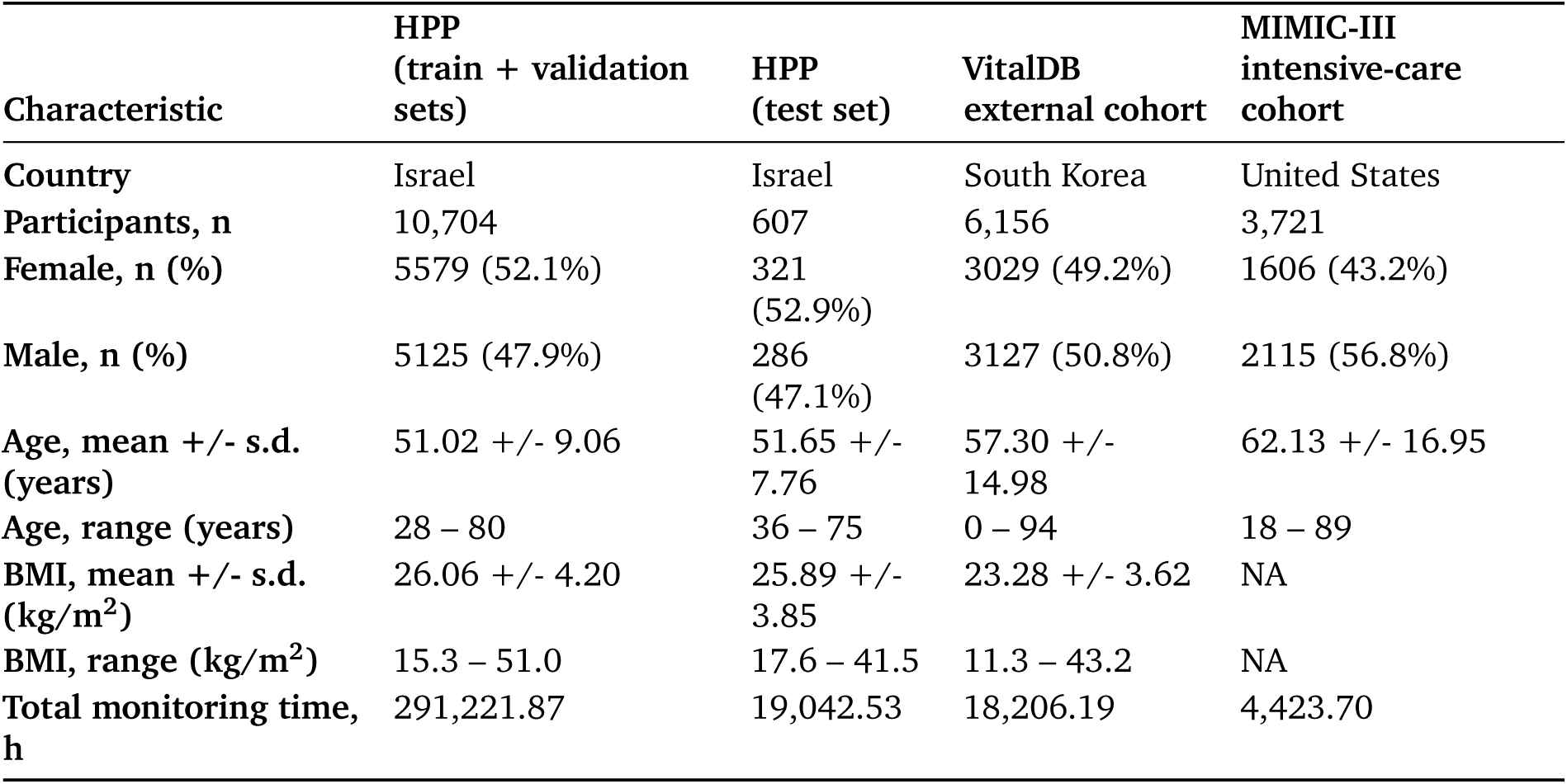
Cohorts Description.

| Characteristic | HPP<br>(train + validation<br>sets) | HPP<br>(test set) | VitalDB<br>external cohort | MIMIC-III<br>intensive-care<br>cohort |
| --- | --- | --- | --- | --- |
| Country | Israel | Israel | South Korea | United States |
| Participants, n | 10,704 | 607 | 6,156 | 3,721 |
| Female, n (%) | 5579 (52.1%) | 321<br>(52.9%) | 3029 (49.2%) | 1606 (43.2%) |
| Male, n (%) | 5125 (47.9%) | 286<br>(47.1%) | 3127 (50.8%) | 2115 (56.8%) |
| Age, mean +/- s.d.<br>(years) | 51.02 +/- 9.06 | 51.65 +/-<br>7.76 | 57.30 +/-<br>14.98 | 62.13 +/- 16.95 |
| Age, range (years) | 28 – 80 | 36 – 75 | 0 – 94 | 18 – 89 |
| BMI, mean +/- s.d.<br>(kg/m<sup>2</sup>) | 26.06 +/- 4.20 | 25.89 +/-<br>3.85 | 23.28 +/- 3.62 | NA |
| BMI, range (kg/m<sup>2</sup>) | 15.3 – 51.0 | 17.6 – 41.5 | 11.3 – 43.2 | NA |
| Total monitoring time,<br>h | 291,221.87 | 19,042.53 | 18,206.19 | 4,423.70 |

Cohort characteristics for the HPP training and validation set, the held-out HPP test set, the VitalDB surgical cohort and the MIMIC-III intensive-care cohort. HPP counts follow one convention: 11,311 participants enrolled (10,704 in the analysis cohort and 607 in the held-out test set, of whom 606 have a valid age prediction). MIMIC-III values cover the 3,721 patients with a recording made within the first day of the first admission. Age is summarised over the 3,522 patients with an age label, since MIMIC-III date-shifts patients older than 89 years; BMI is not available for MIMIC-III. NA, not available.

### Age as a biological-aging anchor

Chronological age serves as an ideal benchmark for evaluating learned physiological representations: it integrates decades of cumulative vascular, metabolic, and autonomic change into a single continuous label, is universally available without additional clinical testing, and is known ^25,26^ to be reflected in peripheral pulse waveform morphology through mechanisms including arterial stiffening and reduced baroreflex sensitivity. Accurate age prediction from raw pulse-oximetry waveforms suggests that the representation captures age-related vascular and autonomic structure, although age may also reflect correlated factors such as medication use, heart rate, and cohort composition. We selected two primary open-source comparators, spanning the two dominant approaches to PPG analysis. PyPPG ^27^ provides a standardized, reproducible implementation of classical PPG fiducial detection and waveform biomarker extraction. PyPPG relies on automated heartbeat segmentation to compute 74 predefined morphological biomarkers, including dicrotic notch position, systolic-to-diastolic ratios, and pulse metrics, derived from fiducial points, and has been validated on large polysomnography-derived PPG datasets, making it an appropriate representative of handcrafted morphology-based PPG analysis. PaPaGei ^11^ is, to our knowledge, the first openly released foundation model for PPG, it is self-supervised on over 2 million unlabelled PPG segments (approximately 57,000 hours) drawn exclusively from publicly available datasets, making it the natural representative of learned rather than engineered PPG representations; we used its released PaPaGei-S model.

Despite this, PulseOx-FM outperformed both comparators on the HPP cohort. On the pretraining set, PulseOx-FM reached *r* = 0.79, compared with *r* = 0.76 for PaPaGei-S and *r* = 0.74 for PyPPG biomarkers, and *r* = 0.63 for features derived from the WatchPAT home sleep-apnea device (Fig. 2a,c, Supp. Table 3), with mean absolute errors of 3.9, 4.2, 4.3 and 5.0 years, respectively, and a negligible mean bias against chronological age (−0.01 years; 95% limits of agreement −9.6 to +9.6 years). On the held-out HPP test set, PulseOx-FM again exceeded PyPPG (*r* = 0.73 vs *r* = 0.69), while PaPaGei-S performed comparably (*r* = 0.73; paired bootstrap *P* = 0.54) (Supp. Table 3). 2D visualization of the latent space using Uniform Manifold Approximation and Projection (UMAP) ^28^ revealed a smooth, continuous gradient along the age and heart rate axes (Fig. 2e–f), indicating that age-related physiology is geometrically organized within the embedding space. A joint projection of the three cohorts placed HPP, VitalDB and MIMIC-III recordings in largely separate regions for both PulseOx-FM and PaPaGei-S, reflecting the cohort shift, while age and heart-rate gradients persisted within each region (Supp. Fig. 2). That a self-supervised model with no explicit waveform engineering outperforms a pipeline specifically designed to extract physiologically meaningful PPG features suggests it has discovered representations that go beyond what manual biomarker design captures.

**Fig. 2.**
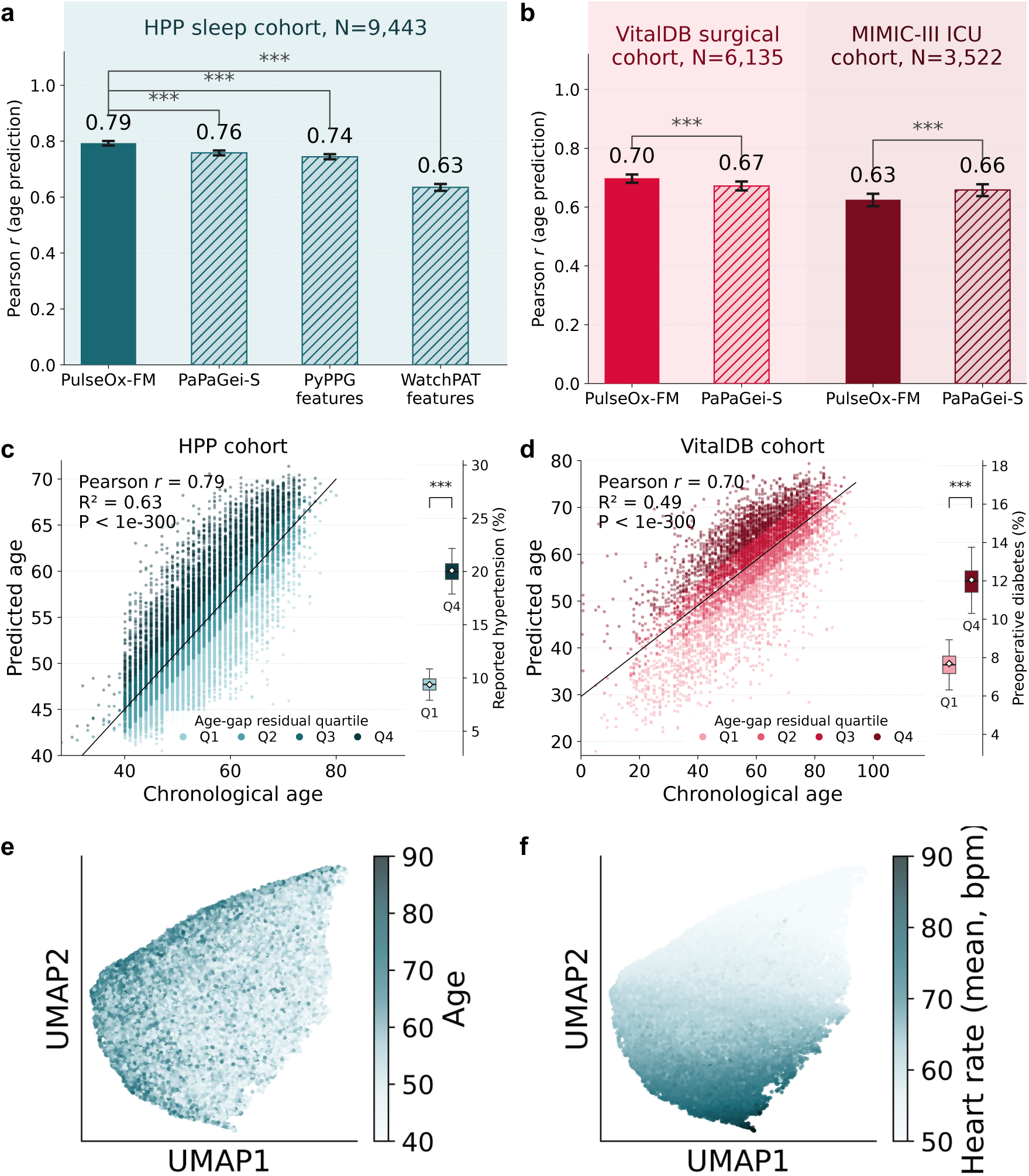
Age prediction as a benchmark for global health encoding, across three cohorts. **a**, Age prediction in the HPP sleep cohort (Pearson *r* between true and predicted age, five-fold crossvalidation) for PulseOx-FM embeddings (solid), PaPaGei-S embeddings, PyPPG features and WatchPAT features (hatched). Bars show the mean *r* across three cross-validation seeds, and error bars the 95% confidence interval from 2,000 participant-clustered bootstrap resamples. Brackets mark the paired difference between PulseOx-FM and each comparator, tested on the same resamples by a two-sided bootstrap test. **b**, The same comparison in the two external cohorts, applied without fine-tuning: the VitalDB surgical cohort and the MIMIC-III intensive-care cohort. Brackets mark only that the two bars differ; which is higher is read from the bars. PaPaGei-S was pretrained on VitalDB and MIMIC-III, so both external cohorts are in-distribution for it but not for PulseOx-FM. Asterisks: ∗∗∗*P* < 0.001. **c, d**, Predicted versus chronological age from PulseOx-FM embeddings, on the same rows as **a** and **b**: HPP (**c**) and VitalDB (**d**). Points are coloured by quartile of the age-gap residual (the residual of predicted age regressed on chronological age), from Q1 (most decelerated, palest) to Q4 (most accelerated, darkest). The black line is the least-squares fit; Pearson *r*, R^2^ and *P* are inset. The strip on the right of each panel gives the prevalence of the strongest accelerated-ageing-associated diagnosis in Q1 against Q4: self-reported hypertension at the same visit in HPP and preoperative diabetes in VitalDB. Boxes summarise 2,000 bootstrap replicates, resampling participants so that each participant’s visits stay together, and show the median, interquartile range and 2.5th–97.5th percentiles; the white diamond is the point estimate. The bracket gives the two-sided bootstrap *P* for the Q4-minus-Q1 difference (∗∗∗*P* < 0.001). **e, f**, Two-dimensional Uniform Manifold Approximation and Projection (UMAP) of the HPP recordings’ PulseOx-FM embeddings, coloured by chronological age (**e**) and by mean heart rate during sleep (**f**).

Ablation experiments confirmed that this performance is specifically attributable to the selfsupervised pretraining objective rather than model architecture alone (Extended Data Fig. 2, Supp. Fig. 3). A baseline model using Fourier-based signal processing could reconstruct the periodic structure of the PPG waveform (Extended Data Fig. 1) but failed to encode information predictive of age (Extended Data Fig. 2b), demonstrating that waveform periodicity and physiological aging are distinct properties. Performance further scaled with segment duration, with 120-second windows providing substantially richer physiological context than shorter segments (Extended Data Fig. 2d–f).

Temporal analysis across the overnight monitoring period, in which segment embeddings are mean-pooled into a recording-level representation (Extended Data Fig. 3), revealed that predictive accuracy was stable throughout the night, with single-segment performance reaching near-plateau levels within the first hour and remaining consistent thereafter (Extended Data Fig. 4a). Notably, pooling segments cumulatively across the night progressively closed the gap with the full-night reference, converging toward ceiling performance by approximately 2–3 hours of monitoring, suggesting that while any single segment captures a meaningful individual-level signal, aggregating even a modest number of early segments substantially recovers the accuracy achievable from a full night. This stability and rapid saturation have an important implication: if the model were learning sleep-stage-specific patterns, rapid-eye-movement architecture, slow-wave activity, arousal frequency, performance would be expected to vary systematically across the night as sleep architecture evolves. The invariance of single-segment accuracy instead suggests that PulseOx-FM is capturing a time-stable, individual-level physiological signature that is expressed consistently regardless of the specific hour of recording. Practically, this means that even a partial night of monitoring, particularly when multiple short segments can be pooled, may be sufficient for meaningful phenotyping. In the external cohorts, pooling the first 20 minutes of intraoperative monitoring in VitalDB (*r* = 0.71, against 0.70 with all data) or the first hour of intensive-care monitoring in MIMIC-III (*r* = 0.62, against 0.63 with all data within the first 24 h from admission) likewise recovered most of the achievable accuracy (Extended Data Fig. 4b,c).

This interpretation is reinforced by the model’s behavior on the VitalDB surgical cohort (N = 6,153), an out-of-distribution dataset (Table 1) recorded under general anesthesia during intraoperative monitoring, a context that differs from the training data in every meaningful respect: patient state, hemodynamic regulation, recording environment, and clinical purpose. PulseOx-FM achieved *r* = 0.70 on this cohort, again exceeding PaPaGei-S (*r* = 0.67), a level of performance that closely approached the held-out HPP test set results (Fig. 2b, d). Notably, VitalDB forms part of PaPaGei’s public pretraining corpus, so this cohort is in-distribution for PaPaGei-S yet out-of-distribution for PulseOx-FM, making the comparison conservative. The fact that embeddings trained exclusively on ambulatory sleep recordings transfer to anesthetized surgical patients without any fine-tuning is strong evidence that PulseOx-FM has learned intrinsic, systemic properties of human vascular physiology rather than artifacts of the sleep context in which it was trained. In the MIMIC-III intensive-care cohort (n = 3,522), PulseOx-FM predicted age with *r* = 0.63 (mean absolute error 10.6 years), below PaPaGei-S (*r* = 0.66; paired difference −0.03, 95% CI −0.05 to −0.02), whose pretraining corpus includes MIMIC-III (Fig. 2b; Supp. Table 3). The PulseOx-age gap, the discrepancy between model-estimated and chronological age corrected for its dependence on chronological age, was also significantly associated with cardiometabolic health: compared with age-matched decelerated agers, accelerated agers had most notably higher blood pressure and more frequent use of beta-blockers (Extended Data Fig. 5; Methods).

### PulseOx-FM embeddings encode broad current health status beyond demographics

A key question for any physiological foundation model is whether its representations carry information about a person’s current health state, beyond what is already explained by basic demographics including age, sex and body mass index (BMI). To test this, we evaluated PulseOx-FM embeddings as predictors of 71 phenotypic targets spanning 52 continuous measures (anthropometrics, blood tests, sleep parameters, dietary intake, imaging-derived traits) and binary outcomes (13 current diagnoses and 6 medication classes intake; Supp. tables 1-2), comparing a model with embeddings added to age, sex, and BMI against demographics alone. All reported improvements were statistically significant after correction for multiple comparisons (Fig. 3, Supp. Table 4).

**Fig. 3.**
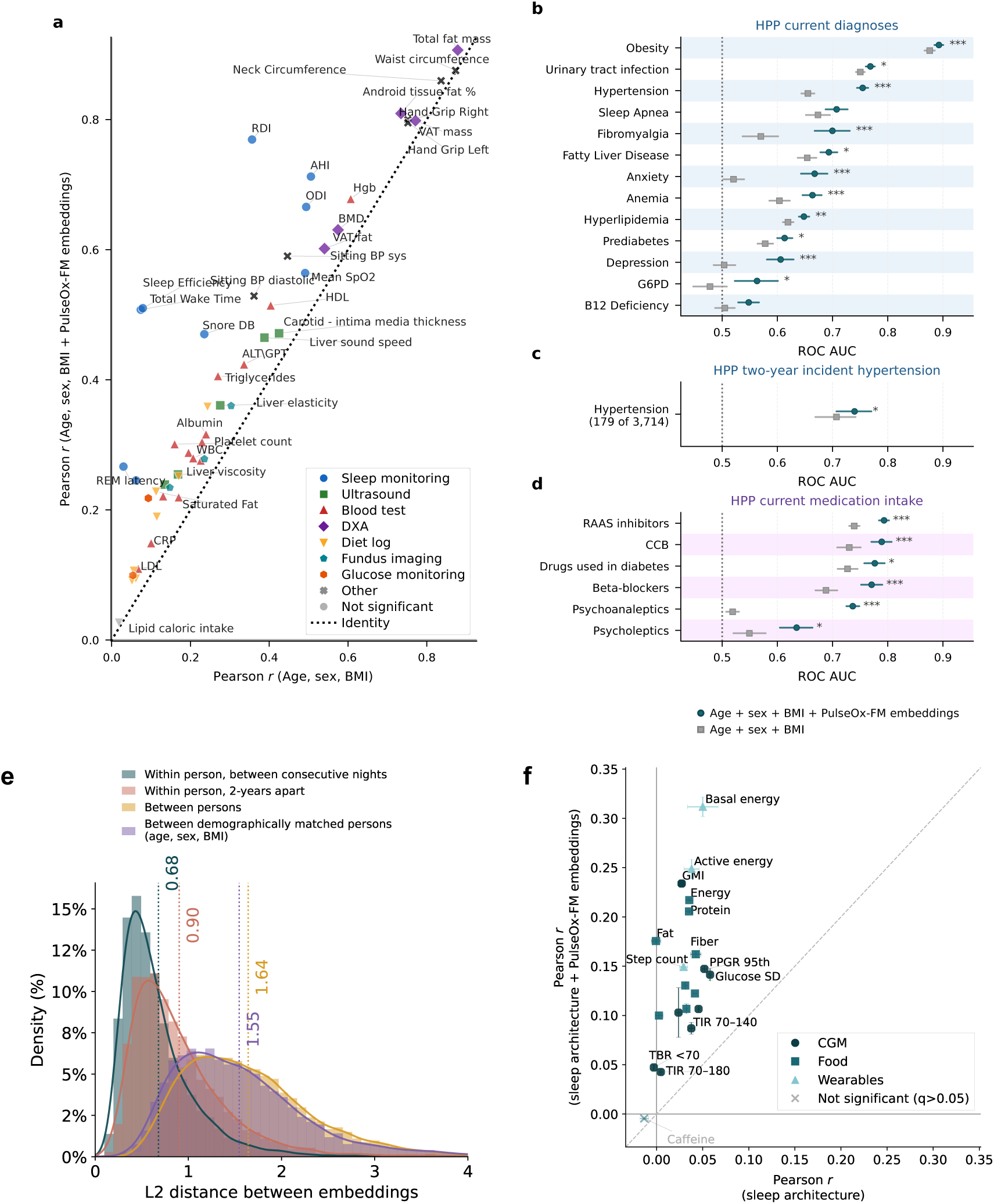
Downstream predictions of multimorbidity-related targets, and the stability of the nocturnal signature. **a,** Prediction performance (Pearson *r* between measured and predicted) for 53 HPP health measures spanning blood tests, dietary logs, sleep monitoring, DXA body composition, US, fundus imaging and continuous glucose monitoring (CGM), for a demographic model (age, sex and body mass index (BMI); horizontal axis) against demographics plus pooled PulseOx-FM embeddings (vertical axis). Targets significantly improved by the embeddings (Benjamini–Hochberg false discovery rate (FDR) *q* < 0.10) are coloured and lie above the dotted identity line; grey circles are non-significant. **b, d**, Receiver operating characteristic area under the curve (ROC AUC) for current self-reported diagnoses (**b**; 13 targets) and current medication intake (**d**; 6 targets) in HPP, for demographics plus PulseOx-FM embeddings (circles) and demographics alone (squares). Horizontal bars are 95% bootstrap confidence intervals (2,000 resamples), the dotted line is chance, and asterisks give the FDR *q* value of the improvement (∗*q* < 0.05, ∗∗*q* < 0.01, ∗∗∗*q* < 0.001). **c,** ROC AUC for incident hypertension within two years in participants normotensive and untreated at baseline (incident: blood pressure ≥ 140/90 mmHg at follow-up, new antihypertensive treatment or a reported diagnosis; n = 3,714; 179 cases). **e,** Distributions of L2 distances between pooled nightly PulseOx-FM embeddings of recordings from the same participant on consecutive nights, from the same participant two years apart, from different participants, and from demographically matched different participants (age, sex and BMI); dotted lines and numbers are medians. **f,** Next-day target prediction (Pearson *r*, cross-validation) from sleep architecture alone (horizontal axis) against sleep architecture plus PulseOx-FM embeddings (vertical axis), by domain: CGM (circles), dietary logging (squares) and wearable-derived activity (triangles). Coloured markers are significantly improved (FDR *q* < 0.05) and grey crosses non-significant. Error bars are the s.d. of *r* across cross-validation seeds. Abbreviations: ALT, alanine aminotransferase; AST, aspartate aminotransferase; BMD, bone mineral density; BP, blood pressure; CCB, calcium channel blockers; CRP, C-reactive protein; eGFR, estimated glomerular filtration rate; G6PD, glucose-6-phosphate dehydrogenase deficiency; GMI, glucose management indicator; Hgb, haemoglobin; HDL/LDL, high-/low-density lipoprotein; PPGR, postprandial glucose response; RAAS, renin–angiotensin–aldosterone system; REM, rapid eye movement; SD, standard deviation; TBR, time below range; TIR, time in range; VAT, visceral adipose tissue; WBC, white blood cell count.

Across continuous measures, embeddings improved predictive performance consistently and significantly over demographics. The largest absolute gains were concentrated in cardiometabolic and vascular domains: systolic blood pressure (0.45 → 0.59, *q <* 0.001), diastolic blood pressure (0.36 → 0.53, *q* < 0.001), carotid intima-media thickness (0.43 → 0.47, *q <* 0.001), triglycerides (0.27 → 0.41, *q* < 0.001), and liver sound speed (0.39 → 0.46, *q <* 0.001). Larger gains still were seen for the sleep-apnoea indices, respiratory disturbance index (RDI, *r*: 0.36 → 0.77, *q* < 0.001) and AHI (0.51 → 0.71, *q* < 0.001); because these and the other sleep parameters are outputs of the WatchPAT device algorithm, derived from the same signal as the embeddings and of limited accuracy as reference labels, these gains show that the representation preserves the information the device extracts rather than providing independent evidence of physiological inference. Body composition traits already well-explained by demographics saw more modest but still significant gains: total fat mass (0.88 → 0.91, *q* < 0.001), android tissue fat percent (0.73 → 0.81, *q <* 0.001), and waist circumference (0.87 → 0.88, *q* < 0.001), suggesting the embeddings contribute information orthogonal to BMI even for anthropometric targets. Notably, several sleep parameters that demographics alone could not predict at all, such as sleep efficiency (*r*: 0.08 → 0.51, *q <* 0.001), total wake time (0.07 → 0.51, *q <* 0.001), and REM latency (0.03 → 0.27, *q* < 0.001), were substantially recovered by the embeddings, consistent with the model having internalized individual-level sleep architecture from the raw waveform. In contrast, dietary intake variables remained the weakest predicted targets even after incorporating embeddings, with daily carbohydrate intake (0.06 → 0.11, *q <* 0.001), daily lipid intake (0.02 → 0.03, *q* = 0.48, the only continuous measure not significantly improved), and daily sodium intake (0.06 → 0.09, *q* < 0.01) showing only marginal absolute performance, suggesting that physiological waveforms recorded during sleep carry limited information about habitual dietary behavior. Similarly, circulating lipid markers such as LDL cholesterol (0.07 → 0.11, *q* < 0.05) and total cholesterol (0.13 → 0.22, *q* < 0.001), as well as renal function as captured by eGFR (0.17 → 0.22, *q <* 0.001), remained relatively weakly predicted, pointing to domains where sleep-derived representations provide only modest additional signal beyond demographics.

Among current diagnoses, the embeddings most strongly identified hypertension (AUC: 0.66 → 0.75, *q* < 0.001) and anxiety (0.52 → 0.67, *q* < 0.001), with significant improvements also observed for fibromyalgia (0.57 → 0.70, *q* < 0.001), anemia (0.60 → 0.66, *q* < 0.001), and depression (0.50 → 0.61, *q* < 0.001) (Fig. 3b). The pattern of the strongest associations clustering around cardiovascular, haematological, and neuropsychiatric conditions is consistent with these systems being the ones most directly reflected in peripheral vascular tone and autonomic modulation, the physiological substrate captured by PPG.

For current medication intake, the embeddings showed their largest gains for psychoanaleptics, antidepressants and stimulants, where demographics alone were essentially uninformative (AUC: 0.52) and embeddings raised performance to 0.74 (*q* < 0.001), a delta of +0.22 that represents the single largest improvement across any classification target. Agents acting on the renin-angiotensin system (0.74 → 0.79, *q* < 0.001), beta-adrenergic blocking agents (0.69 → 0.77, *q* < 0.001), and calcium channel blockers (0.73 → 0.79, *q* < 0.001) all showed significant gains, forming a coherent cluster of antihypertensive drug classes whose detection from PPG is mechanistically plausible given their direct cardiovascular effects (Fig. 3d). That three independent classes of antihypertensive medication are all detectable above demographics suggests the embeddings are not simply recovering a proxy for blood pressure, but are encoding broader pharmacodynamic signatures of these agents in the peripheral vasculature.

Further evidence of generalization came from the VitalDB surgical cohort, where embeddings derived from intraoperative PPG recordings improved prediction of preoperative hemoglobin (r: 0.46 → 0.52, *q* < 0.001) and preoperative glucose (0.21 → 0.23, *q* = 0.03), and contributed to identification of preoperative diabetes (AUC: 0.65 → 0.67, *q* = 0.048) (Fig. 4a). While the absolute gains are more modest than those observed in the HPP cohort, expected given the domain shift to an acute surgical context, the capacity of intraoperative embeddings to recover preoperative clinical state provides additional evidence that PulseOx-FM encodes stable, individual-level physiological signatures that persist across recording contexts.

**Fig. 4.**
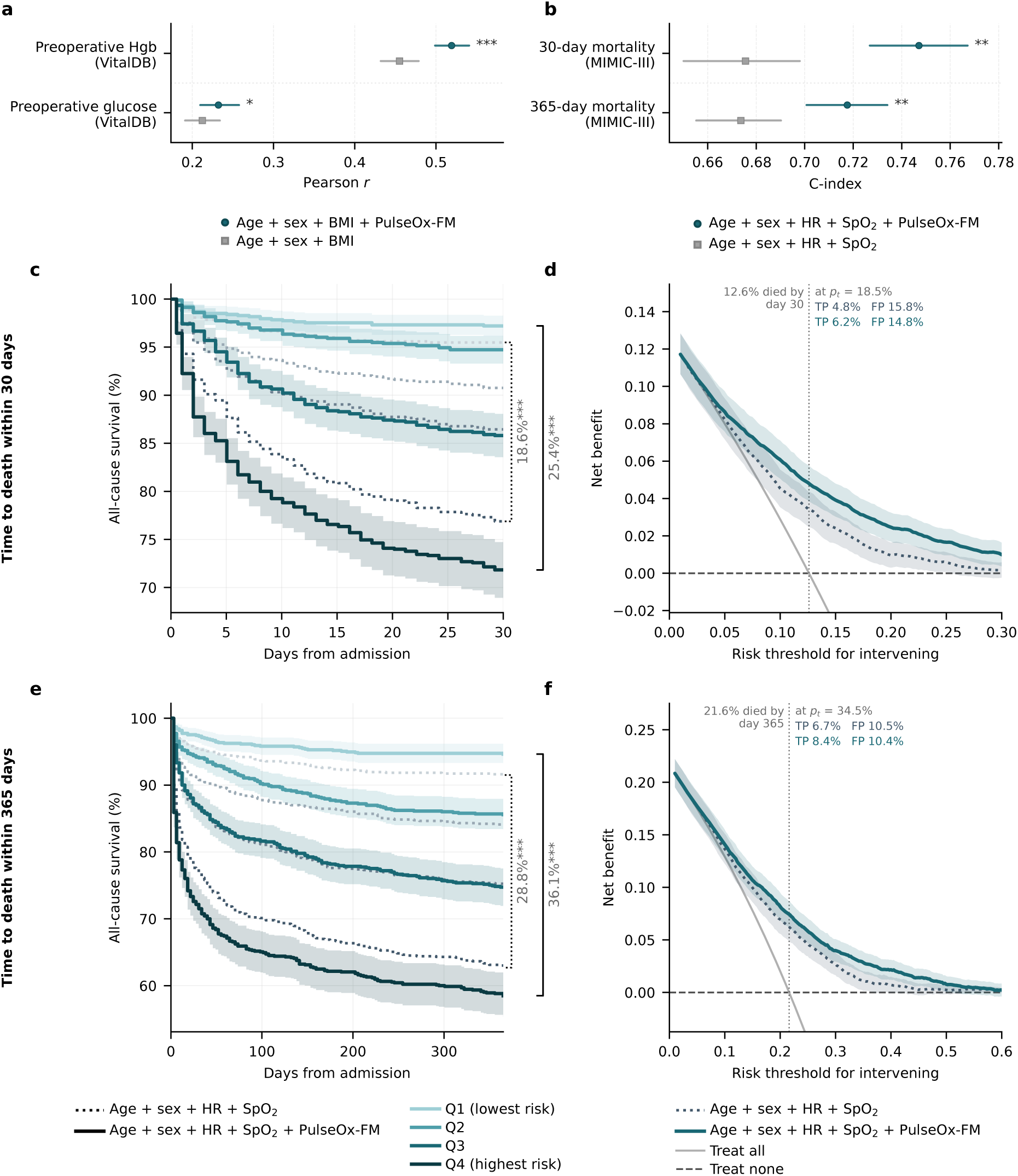
Transfer to two external cohorts: preoperative measures in VitalDB and mortality risk in MIMIC-III, read as discrimination and decision-analytic benefit. **a,** Pearson *r* for preoperative haemoglobin (Hgb) and glucose in the VitalDB surgical cohort, for age, sex and BMI plus PulseOx-FM embeddings (circles) and for age, sex and BMI alone (squares). Bars are 95% bootstrap confidence intervals (2,000 resamples); asterisks give the FDR *q* value of the improvement (∗*q* < 0.05, ∗∗∗*q* < 0.001). **b,** Harrell’s concordance index (C-index) for 30-day and 365-day all-cause mortality in the MIMICIII intensive-care cohort (n = 3,721 patients; 469 and 805 deaths), for Cox models on age, sex, mean heart rate and mean oxygen saturation plus PulseOx-FM embeddings (circles) and on those four covariates alone (squares; BMI is not available in MIMIC-III). Heart rate and saturation are the vital signs the pulse oximeter itself reports over the same 120-s windows the encoder read. Bars are 95% confidence intervals from 1,000 patient-level bootstrap resamples; asterisks give the two-sided paired bootstrap *P* for the increment (*P* = 0.002 at both horizons, the resolution floor; ∗∗). **c, e**, All-cause survival by quartile of out-of-fold predicted Cox risk at 30 days (**c**) and 365 days (**e**) (about 930 patients per quartile), for age, sex, heart rate and oxygen saturation alone (dotted) and with PulseOx-FM added (solid); lightness runs from Q1 (lowest risk) to Q4 (highest risk). Curves are Kaplan–Meier estimates, and the shaded bands, drawn for the PulseOx-FM curves only, are 95% Greenwood intervals. Brackets give the Q1-minus-Q4 separation in survival at the horizon. Asterisks give a log-rank test of each model’s own Q1 against its Q4, Benjamini–Hochberg-adjusted across models and horizons (∗∗∗*q* < 0.001). **d, f**, Decision curves for the same models at 30 days (**d**) and 365 days (**f**): net benefit against the risk threshold for intervening, with the treat-all and treat-none strategies for reference. The dotted vertical line marks the observed event rate. Curves are the mean over three cross-validation seeds, and bands are 95% patient-level bootstrap intervals (2,000 resamples). FP, false positives; HR, heart rate; SpO_2_, peripheral oxygen saturation; TP, true positives.

### Early risk stratification in intensive care

We then asked whether the representation can inform a clinical decision, using an input available when the decision is made and the comparator a clinician has at that point. We applied the frozen encoder, without fine-tuning, to routine bedside pulse oximetry from the first day of admission in the MIMIC-III intensive-care cohort (n = 3,721; 469 deaths by 30 days and 805 by 365 days; Methods). The comparator is what the same device already provides: age and sex together with the mean heart rate and oxygen saturation measured over the identical recording windows the encoder read, these being the vital signs available wherever such a model would be applied. Adding the pooled embeddings to that baseline raised the concordance index (C-index) for 30-day all-cause mortality from 0.676 (95% CI 0.650–0.698) to 0.747 (0.727– 0.767), a gain of +0.072 (+0.051 to +0.092), and for 365-day mortality from 0.674 (0.655–0.690) to 0.718 (0.701–0.734) (+0.044, +0.030 to +0.057) (Fig. 4b); the embeddings alone reached 0.722 at 30 days. Stratified by quartile of out-of-fold predicted risk, the survival gap between the lowest- and highest-risk quartiles widened, when the embeddings were added, from 18.6 to 25.4 percentage points at 30 days and from 28.8 to 36.1 points at 365 days, with risk refined in both directions (Fig. 4c,e). In decision-curve analysis, adding the embeddings increased net benefit across the range of thresholds at which a clinician might act: at a 10% threshold for 30-day death, the gain was equivalent to 13.9 fewer unnecessary interventions per 100 patients (95% CI 9.5–18.1) for the same number of deaths correctly flagged (Fig. 4d,f). At that threshold the model with the embeddings flags 43.9 patients per 100 and catches 9.9 of the 12.6 deaths among them (78.3% of all deaths), against 53.4 flagged and 9.4 deaths caught (74.8%) on the clinical covariates alone. With discharge alive treated as a competing event, the cumulative incidence of in-hospital death by day 28 was 1.8% in the lowest and 23.2% in the highest predicted-risk quartile (Supp. Fig. 4b). Against PaPaGei-S, whose pretraining corpus includes MIMIC-III, PulseOx-FM discriminated in-hospital mortality more accurately (AUROC 0.757 vs 0.731; difference +0.026, 95% CI +0.005 to +0.047), whereas the differences in all-cause survival discrimination between the two encoders were not distinguishable from zero on this cohort (Supp. Fig. 4a,c,d; Supp. Table 4).

### Longitudinal trajectories

We next asked whether overnight PulseOx-FM representation contains information about future vascular risk. PulseOx-FM embeddings predicted the two-year incidence of hypertension in participants who were normotensive and untreated at the time of recording (AUC: 0.71 → 0.74, embeddings only: 0.69, Fig. 3c), suggesting that the vascular signatures encoded in a single overnight oximetry recording carry forward-looking information about cardiovascular disease trajectory, not merely a reflection of current disease burden. The gain rests on 179 incident cases among 3,714 participants and is modest (increment +0.03, 95% CI +0.002 to +0.065; paired bootstrap *P* = 0.039). When baseline systolic and diastolic blood pressure were added to the comparator, the embeddings improved discrimination only marginally (difference in AUROC +0.017, 95% CI +0.002 to +0.034; DeLong *P* = 0.03) and did not increase net benefit at risk thresholds of 10–20% (between −0.35 and +0.26 unnecessary interventions avoided per 100 people screened). We therefore report this result as an association and do not put it forward as a clinical application.

As part of the HPP study design, participants wore continuous glucose monitors (CGM), logged dietary intake, and carried activity trackers in parallel with overnight pulse oximetry, providing a rare opportunity to test whether nightly embeddings track day-to-day fluctuations in metabolic state within the same individual. To quantify the temporal structure of the embedding space, we computed L2 distances between embeddings under four conditions: within the same person between consecutive nights, within the same person two years apart, between different persons matched on age, sex, and BMI, and between unmatched persons (Fig. 3e). The resulting distributions were clearly ordered: consecutive-night within-person distances were the smallest (median L2: 0.68), followed by within-person distances two years apart (0.90), demographically matched between-person distances (1.55), and unmatched between-person distances (1.64). This hierarchy, where the same individual separated by two years is still more similar to themselves than to a demographically matched stranger, demonstrates that PulseOx-FM embeddings encode a stable, individual-specific physiological fingerprint that persists over time while remaining sensitive to longitudinal change. This individual specificity extended to next-day metabolic targets: the absolute difference between predicted next-day GMI, basal energy expenditure, and dietary energy intake for pairs of recordings was markedly smaller within the same participant across consecutive nights than between participants (Extended Data Fig. 6). This confirms that the model is tracking genuine intra-individual metabolic variation rather than simply recovering population-level demographic patterns.

To test whether this within-person signal carries information beyond the sleep metrics already derivable from consumer devices, we benchmarked the embeddings against a sleep-architecture baseline rather than against static demographics. Sleep architecture, including total sleep duration and the proportion of light, deep (slow-wave), and REM sleep, was chosen as the comparator for two reasons. First, unlike age, sex, and BMI, which are fixed across the recording, sleep architecture varies appreciably from night to night within the same individual ^29,30^, making it a far stricter baseline for any claim of a night-specific predictive signal. Second, these are precisely the metrics that smartwatches and other consumer wearables compute and surface to estimate next-day readiness and recovery ^31^, so improvement over them speaks directly to added value beyond commercially available sleep tracking. Comparing next-day prediction from sleep architecture alone against sleep architecture combined with PulseOx-FM embeddings (Fig. 3f, Supp. Table 5), the embeddings significantly improved 21 of 22 next-day targets, spanning continuous glucose monitoring, dietary logging, and wearable-derived activity. Sleep architecture alone was nearly uninformative (Pearson r ≈ 0.03–0.06 for most targets), whereas adding the embeddings raised performance several-fold: next-day glucose management indicator (GMI, r: 0.03 → 0.23), dietary energy intake (0.04 → 0.22), basal energy expenditure (0.05 → 0.31), active energy expenditure (0.04 → 0.25), and step count (0.03 → 0.15); only next-day caffeine intake was not significantly improved. That the embeddings add significant next-day predictive signal over the very sleep-stage features wearables already compute, and over a baseline that itself carries genuine night-to-night variation, demonstrates that PulseOx-FM captures waveform-level physiology not reducible to sleep-stage composition.

### A direct, diet-independent next-day glycemic signal

To probe the next-day metabolic associations above more deeply, we used a literature-driven agentic analysis system as the analytical tool. The system pairs a large language model with the HPP data, the PulseOx-FM embeddings, and a literature-search tool to propose literature-grounded hypotheses, each paired with an executable analysis workflow and reviewed by a human before execution (Fig. 5a). Using this framework, we addressed the central question raised by the next-day analyses: is the embedding’s association with next-day glycemia a genuine physiological signal, or merely a shadow of the following day’s diet?

**Fig. 5.**
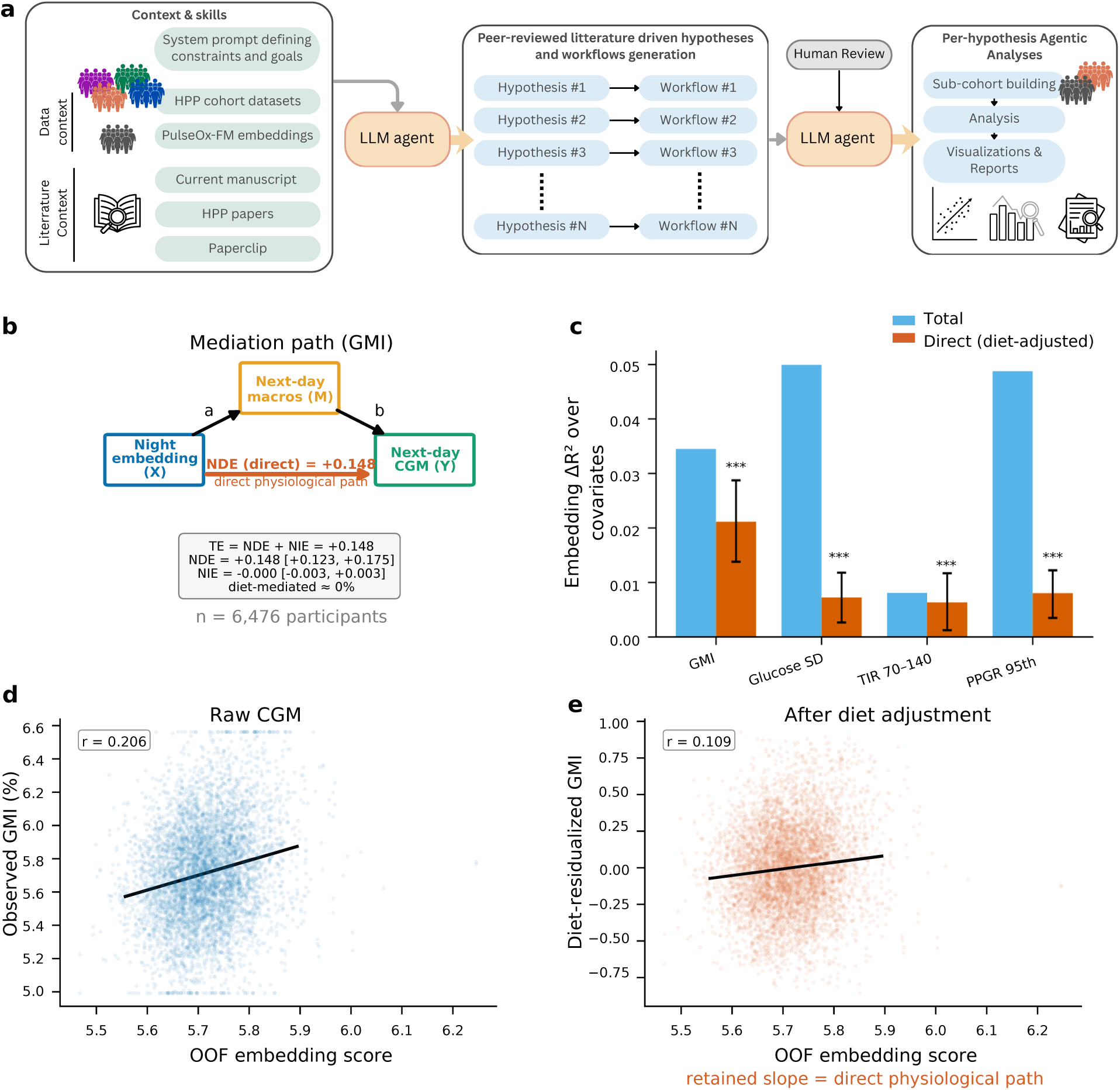
Literature-driven agentic discovery reveals a diet-independent association between sleep physiology and next-day glycaemic control. **a,** Schematic of the agentic analysis pipeline. A large language model (LLM) agent is provided with data context (HPP cohort datasets, PulseOx-FM embeddings, and the current manuscript) and literature context (HPP publications and the *Paperclip* literature-search tool ^49^) and autonomously generates peer-reviewed literature-driven hypotheses paired with executable analysis workflows. Each hypothesis workflow pair undergoes human review before the agent executes per-hypothesis analyses comprising sub-cohort construction, statistical analyses, and visualisations. One of the hypotheses generated and tested by the agent is showcased in **b-e**. **b,** Mediation path diagram for the association between the nightly sleep embedding (X) and next-day GMI (Y), with next-day dietary macronutrient intake (M) as the candidate mediator. The natural indirect effect (NIE) is indistinguishable from zero, while the natural direct effect (NDE) accounts for the full total effect (TE), indicating that the sleep-embedding–glycemia association is not explained by dietary intake. **c**, Embedding `Δ`R^2^ over demographic covariates for four CGM-derived glycemic targets: GMI, glucose SD, TIR 70–140 mg/dL, and PPGR 95th percentile. Blue bars show the total embedding contribution; orange bars show the direct contribution after partialling out next-day macronutrient intake. Error bars indicate 95% bootstrap confidence intervals; significance was determined by two-sided permutation tests with Benjamini–Hochberg FDR correction, and asterisks denote the FDR-adjusted q-value (∗∗∗ *q* < 0.001). **d**, Scatter plot of the out-of-fold (OOF) PulseOx-FM embedding predictive score versus observed next-day GMI in the raw CGM data (*r* = 0.206, n = 6,476 participants). **e,** Scatter plot of the same embedding score versus diet-residualized GMI (GMI with next-day macronutrient intake regressed out), confirming that the association persists after dietary adjustment (*r* = 0.109), consistent with a direct physiological path. CGM, continuous glucose monitoring; `Δ`R^2^, change in coefficient of determination; GMI, glucose management indicator; HPP, Human Phenotype Project; PPGR, postprandial glucose response; SD, standard deviation; TIR 70–140, time in glucose range 70–140 mg/dL.

Applying a causal-mediation framework on 6,476 participants (14,526 person-days, the subcohort with concurrent sleep, dietary and CGM data), we decomposed the effect of the nightly embedding (X) on next-day glycemic control (Y) into a path mediated by next-day macronutrient intake (M) and a direct path (Fig. 5b). Across four CGM-derived targets, GMI, glucose SD, time-in-range 70–140 mg/dL, and 95th-percentile postprandial response, the diet-mediated (natural indirect) effect was indistinguishable from zero (≈0–10%), while the direct (natural direct) path accounted for essentially the entire total effect and remained significant after FDR correction (direct `Δ`R^2^ for GMI +0.021, 95% CI [0.014, 0.029], permutation p = 0.001; Fig. 5c). The association persisted after dietary adjustment: the embedding-derived score correlated with raw next-day GMI (*r* = 0.21) and with diet-residualized GMI (*r* = 0.11) (Fig. 5d–e). A sensitivity analysis confirmed the direct GMI path is robust to adjustment for anti-diabetic medication, visceral adiposity, habitual (rather than sameday) diet, additional dietary mediators (caffeine and ultra-processed food), and dietary-mediator measurement error (Supplementary Figure 5). The discovery is therefore mechanistic, not methodological: the link between overnight pulse-oximetry physiology and next-day glycemic control is predominantly a direct physiological effect, not a downstream consequence of next-day food intake.

## Discussion

Wearable photoplethysmography is among the most scalable physiological sensing technologies available, yet its clinical utility has remained largely confined to conventional vital-sign measurements and monitoring of heart rate and oxygen saturation. Here we introduce PulseOx-FM, a transformer-based foundation model pretrained on large-scale overnight pulse oximetry via masked autoencoding, and demonstrate that self-supervised representations derived from this single wearable-compatible signal support broad clinical phenotyping spanning cardiometabolic risk, disease incidence, medication exposure, and next-day metabolic state. This positions PulseOx-FM at the intersection of two otherwise divergent paradigms: the predictive depth of comprehensive sleep-based physiological modelling ^32^ and the accessibility of consumer-grade wearable sensing, a combination that prior approaches have not achieved simultaneously.

We separate two kinds of claim that these results support. The first is clinical utility, which we restrict to settings in which the input is available at the moment the decision is made, the output takes the form of that decision, and the comparator is the information the clinician actually has. Early risk stratification in intensive care meets all three: the input is the pulse-oximetry waveform already recorded by the bedside monitor within the first day of admission; the output is a mortality risk that can inform level-of-care decisions, such as intensive care versus step-down care, transfer to a higher-level centre and bed allocation under capacity pressure, and that, if tracked over time, could support the detection of deterioration; and the comparator is age, sex and the heart rate and oxygen saturation the same pulse oximeter already reports, all available before laboratory tests or imaging. In this setting, PulseOx-FM, transferred without fine-tuning from healthy sleeping adults to acutely ill patients recorded on hospital monitors in another country, added substantially to those bedside measurements in discrimination, risk stratification and net benefit. The second is information content: the phenotypic target panel, the WatchPAT sleep indices, the preoperative measures in VitalDB and the associations of the PulseOx-age gap show what the representation encodes, but they are association-level evidence, and we attach no clinical claim to them.

As a summary measure of cumulative biological change, chronological age is among the most informative proxies of overall physiological health status. Its imprint on peripheral pulse waveform morphology is well established: arterial stiffening reduces waveform compliance, attenuates the dicrotic notch, and shortens time to peak systolic pressure, while declining baroreflex sensitivity alters beat-to-beat variability in ways reflected in the low-frequency structure of the PPG signal ^25,26,33,34^. PulseOx-FM outperformed both PyPPG and the proprietary WatchPAT feature set and this gap is mechanistically informative. Handcrafted pipelines such as PyPPG and WatchPAT are architecturally optimised for respiratory event detection; their morphological features are most sensitive to the abrupt, breath-coupled oscillations characterising obstructive apnoea, not the slow inter-beat waveform changes associated with vascular ageing that unfold across timescales of minutes to hours. Operating on 120-second windows without any prescribed morphological target, PulseOx-FM captures this longer-range temporal structure. This motivates the concept of a PulseOx-age gap, the discrepancy between chronological age and model-estimated age, as a potential index of accelerated or decelerated vascular ageing, analogous to epigenetic clocks ^35^ ^36^ but derivable from a standard overnight wearable recording.

PulseOx-FM demonstrated predictive signals across a broad phenotypic spectrum, spanning neurological, haematological, respiratory, and metabolic conditions, yet the strongest and most consistent embedding-driven improvements concentrated on cardiometabolic disease. The continuous measures showing the largest absolute improvements: respiratory disturbance index, apnoea-hypopnea index, systolic and diastolic blood pressure, triglycerides, carotid intima-media thickness, and liver sound speed, are individually components, sequelae, or diagnostic correlates of the metabolic syndrome. Blood pressure and triglycerides are defining criteria ^37^; sleep-disordered breathing is a recognised precipitant and consequence of the syndrome ^38^; elevated carotid intima-media thickness is an independent structural marker of the syndrome in both sexes ^39^ and liver steatosis, now reframed as metabolic dysfunction-associated steatotic liver disease, is widely considered the hepatic manifestation of metabolic syndrome ^40,41^. Taken together, these measures represent a cluster of interconnected factors that increase the risk of cardiovascular atherosclerotic diseases and diabetes mellitus type 2 ^37^. Among medication classes, the four with the highest absolute embedding-based predictive performance, renin-angiotensin system agents, calcium channel blockers, drugs used in diabetes and beta-blockers, represent the pharmacological backbone of cardiometabolic management, while hypertension was among the diagnoses most improved by embeddings. This convergence is physiologically coherent: the peripheral vasculature is a primary effector organ for metabolic syndrome pathophysiology, with insulin resistance impairing endothelial function, chronic sympathetic activation elevating vascular tone, and intermittent hypoxia from sleep-disordered breathing driving oxidative stress reflected in peripheral arterial compliance ^38,42^. Nocturnal acquisition further reduces confounding from the postural, behavioural, and emotional variation of waking life. This framing positions PulseOx-FM as a specific digital biomarker for cardiometabolic risk. Its capacity to predict two-year hypertension incidence in normotensive participants suggests that sub-clinical vascular signals detectable before diagnosis are already encoded in the waveform, addressing a major unmet need in preventive cardiology ^43^.

Notably, the model’s sensitivity was not confined to cardiometabolic morbidity. When improvement was instead quantified as the embedding-driven gain over a demographics-only baseline, the single largest increase across all classification targets was for psychoanaleptics, antidepressants and stimulants, a class for which demographics were uninformative. The fact that the discriminative signal stemmed almost exclusively from the waveform aligns with the detection of drug-induced changes in autonomic tone ^44^, extending the model’s pharmacodynamic sensitivity beyond the cardiovascular system.

Beyond cross-sectional phenotyping, nightly PulseOx-FM embeddings tracked within-person next-day metabolic, dietary, and activity state, including glycemic control, energy expenditure, and dietary intake., Critically, benchmarking against a sleep-architecture baseline demonstrated that the embeddings improved next-day prediction for nearly all metabolic, dietary, and activity targets even relative to the light, deep, and REM sleep proportions that consumer wearables already compute, dissociating the predictive signal from sleep-stage composition and implicating subtler waveform-level changes, plausibly in autonomic tone or nocturnal respiratory patterning, as the underlying information source. This is consistent with evidence that nocturnal heart rate variability encodes next-day insulin sensitivity ^45^ and extends it to the raw waveform domain without explicit feature engineering, suggesting a potential pathway toward passive metabolic monitoring using devices already deployed at population scale. We corroborated this dissociation with a general-purpose, literature-driven agentic analysis system, which independently established that the embedding’s effect on next-day glycemic control is largely direct rather than mediated by next-day diet. Beyond this specific result, the agentic framework is general-purpose: it can be directed at any association in the cohort to generate and test literature-grounded hypotheses, and we anticipate it serving as a reusable engine for hypothesis-driven discovery on foundation-model representations more broadly.

The sleep context should be interpreted primarily as a standardized physiological acquisition window rather than as evidence that the learned representation is sleep-specific. Overnight monitoring reduces behavioral, postural, and emotional variation, allowing stable vascular and autonomic signatures to emerge. The VitalDB transfer result further suggests that these signatures are not confined to natural sleep, but reflect generalizable peripheral hemodynamic physiology.

Several features of the study lend confidence to the findings. The masked-reconstruction objective and a standard Vision Transformer were chosen because reconstruction does not depend on the label-preserving augmentations that joint-embedding objectives require, which have no established physiological analogue for coupled multichannel pulse oximetry; the contribution lies in the scale and phenotypic depth of the pretraining corpus, the largest PPG pretraining corpus by recording duration to our knowledge. The pretraining dataset is among the largest assembled for a PPG foundation model, comprising over 310,000 hours from 11,311 deeply phenotyped participants with concurrent biochemistry, body composition imaging, continuous glucose monitoring, actigraphy, and real-time dietary logging, a breadth of concurrent phenotyping rare in physiological modelling studies. Longitudinal follow-up with repeat recordings two years apart further allowed temporal embedding stability to be characterised in a way that cross-sectional designs cannot support. Finally, pulse oximetry’s ubiquity across clinical and consumer settings at a cost and form factor orders of magnitude more accessible than the polysomnographic inputs required by prior comprehensive physiological models ^18^ directly enables the population-scale deployment at which preventive health gains are most meaningful.

Limitations must nonetheless be carefully considered. The HPP cohort is predominantly a relatively healthy Israeli population, which may limit generalisability to populations with higher cardiometabolic disease burden or distinct ethnic vascular phenotypes. The VitalDB transfer experiment partially addresses this concern: the surgical cohort, recruited in South Korea and comprising patients with active comorbidities undergoing general anaesthesia, differs from the HPP sample in ethnicity, clinical acuity, and haemodynamic context, yet PulseOx-FM embeddings recovered chronological age and improved prediction of preoperative haemoglobin and glucose without fine-tuning, supporting at least partial cross-population and cross-clinical-context generalisability. The MIMICIII analysis extends this to older, acutely ill and more ethnically diverse adults recorded on hospital bedside monitors in the United States. Because pulse oximetry is an optical measurement that skin pigmentation is known to affect ^46^, we examined performance across subgroups. In HPP, age prediction was stable across ancestry groups derived from grandparents’ countries of birth, and PulseOx-FM exceeded PaPaGei-S in all seven groups (Supp. Fig. 6a,b). In MIMIC-III, discrimination of 30-day mortality by the embeddings did not differ across self-reported ethnicity and was at least as high in Black or African American as in White patients (Supp. Fig. 6c–f). Ancestry and self-reported ethnicity are proxies for skin pigmentation rather than measurements of it, so these analyses do not establish that finger photoplethysmography, or a model built on it, is insensitive to pigmentation. A further key limitation is the inability to disentangle physiological signatures of disease from those of pharmacological treatment. Several conditions detected by PulseOx-FM, are commonly managed with agents that directly reshape nocturnal cardiovascular and autonomic physiology: beta-blockers enhance heart rate variability, calcium channel blockers and RAAS agents modify peripheral resistance and pulse wave velocity, and psychoanaleptics can perturb autonomic tone and sleep architecture ^47,48^. Strong performance for these conditions and their corresponding medication classes therefore likely reflects a mixture of disease-related and drug-induced signals that cannot be resolved in the current design, a limitation compounded by reliance on self-reported diagnoses and medication use. Future work incorporating objective medication verification and treated-versus-untreated comparisons within the same diagnosis will be needed to partition these contributions. Prospective validation of the PulseOx-FM-derived vascular age against cardiovascular events and mortality will also be required before it can be considered for clinical use. Finally, while UMAP visualisations reveal organised gradients along age and heart rate axes, systematic attribution of embedding dimensions to specific physiological mechanisms requires further work combining activation analysis and concept-based probing.

The clinical-utility analysis also has specific limitations. It is a retrospective, out-of-fold evaluation in a cohort of convenience, and it has not been tested prospectively, which is its main limitation. Before use in intensive care, the model requires a prospective, silent-mode evaluation on live bedside monitors, in which predictions are recorded but not acted upon, recalibration to local event rates (our decision curves fix the average predicted risk to the observed event rate, so calibration in the large must be re-established at each site), and comparison with established severity scores once these become available during the stay. For cardiometabolic use, the model would need validation on the clinical and consumer devices on which it would be deployed, prospective testing against incident outcomes and comparison with standard risk scores at clinically meaningful thresholds. In both settings, performance should be monitored across skin pigmentation and other subgroups.

In conclusion, PulseOx-FM demonstrates that a foundation model pretrained on large-scale overnight pulse oximetry encodes a physiologically rich, individually stable, and contextually transferable representation of cardiovascular and autonomic state. Its predictive reach, spanning biological ageing, cardiometabolic risk, early mortality risk in intensive care, and next-day glycaemic regulation, is achieved from a sensor already deployed in consumer wearable devices at population scale, defining a plausible pathway toward passive, continuous, and accessible longitudinal health monitoring. The convergence of the model’s strongest signals around the metabolic syndrome complex is consistent with this system’s pervasive effects on peripheral vascular tone, endothelial function and autonomic regulation; the precise physiological substrate captured by the PPG waveform. PulseOx-FM establishes that wearable pulse oximetry, interpreted through modern representation learning, can support physiological inference previously reserved for multi-sensor clinical instruments. Nonetheless, translating these results into clinical practice will require prospective validation in diverse populations and resolution of pharmacological confounding.

---

**Extended Data Fig. 1.**
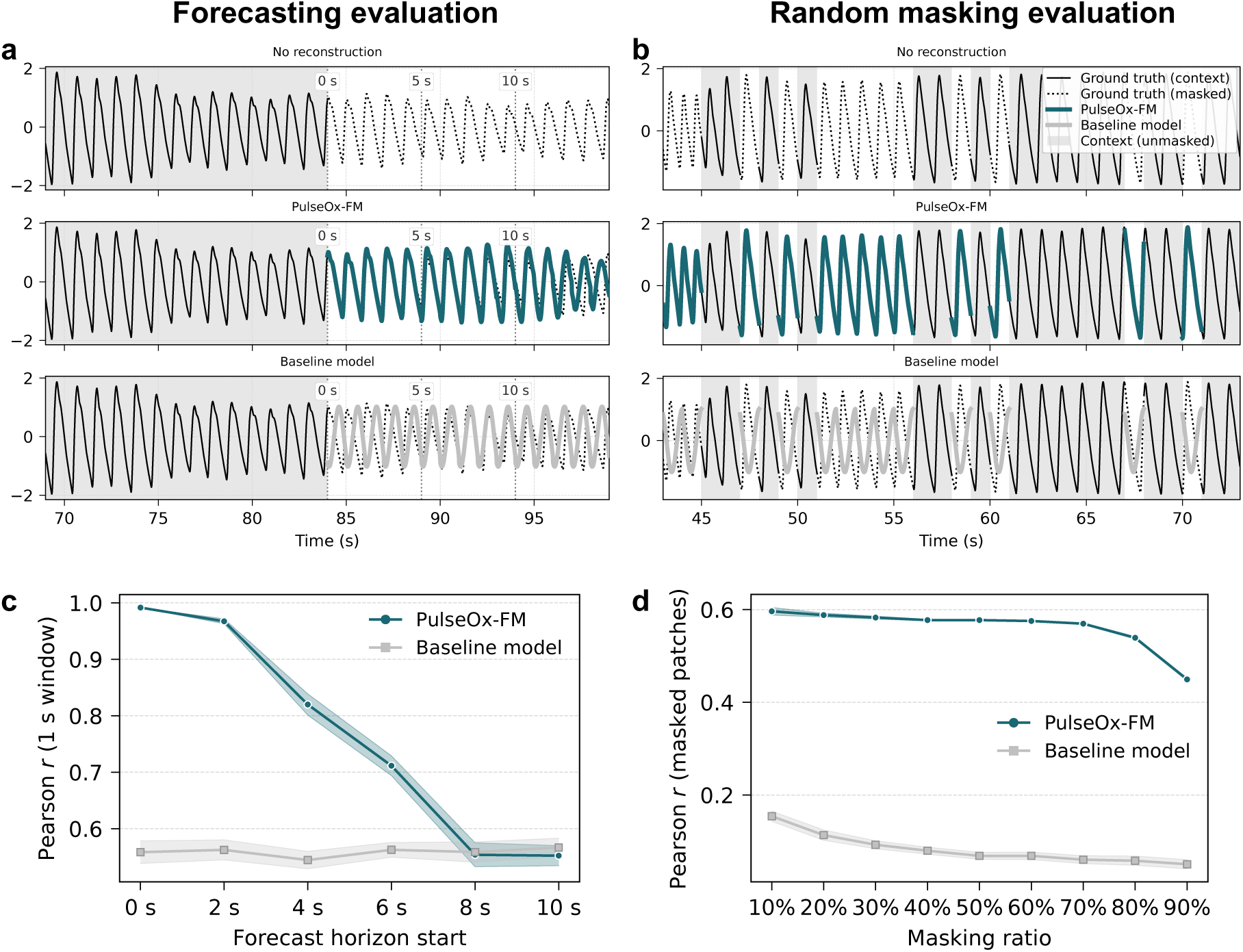
PulseOx-FM performance in physiological signal reconstruction. **a**, Representative example of signal forecasting, from a held-out test recording, showing unmasked context (grey area) and comparing the masked ground truth (black dotted lines), with reconstructions from PulseOx-FM (teal line) and a classical Fourier-based baseline (gray line). **b**, Representative example of a random masking reconstruction from the held-out test subset using the same visual conventions as in a, and demonstrating the model’s ability to reconstruct missing patches of the physiological waveform **c**, Quantitative evaluation of forecasting accuracy (Pearson *r* between ground truth and reconstruction within a 1 second window) across increasing temporal horizons. **d**, Quantitative evaluation of reconstruction (Pearson *r* between ground truth and reconstruction within the masked patches) fidelity across varying masking ratios (10% to 90%). Data is presented as mean values across the 1000 evaluated samples from the held-out test set, with shaded regions representing the 95% confidence intervals.

**Extended Data Fig. 2.**
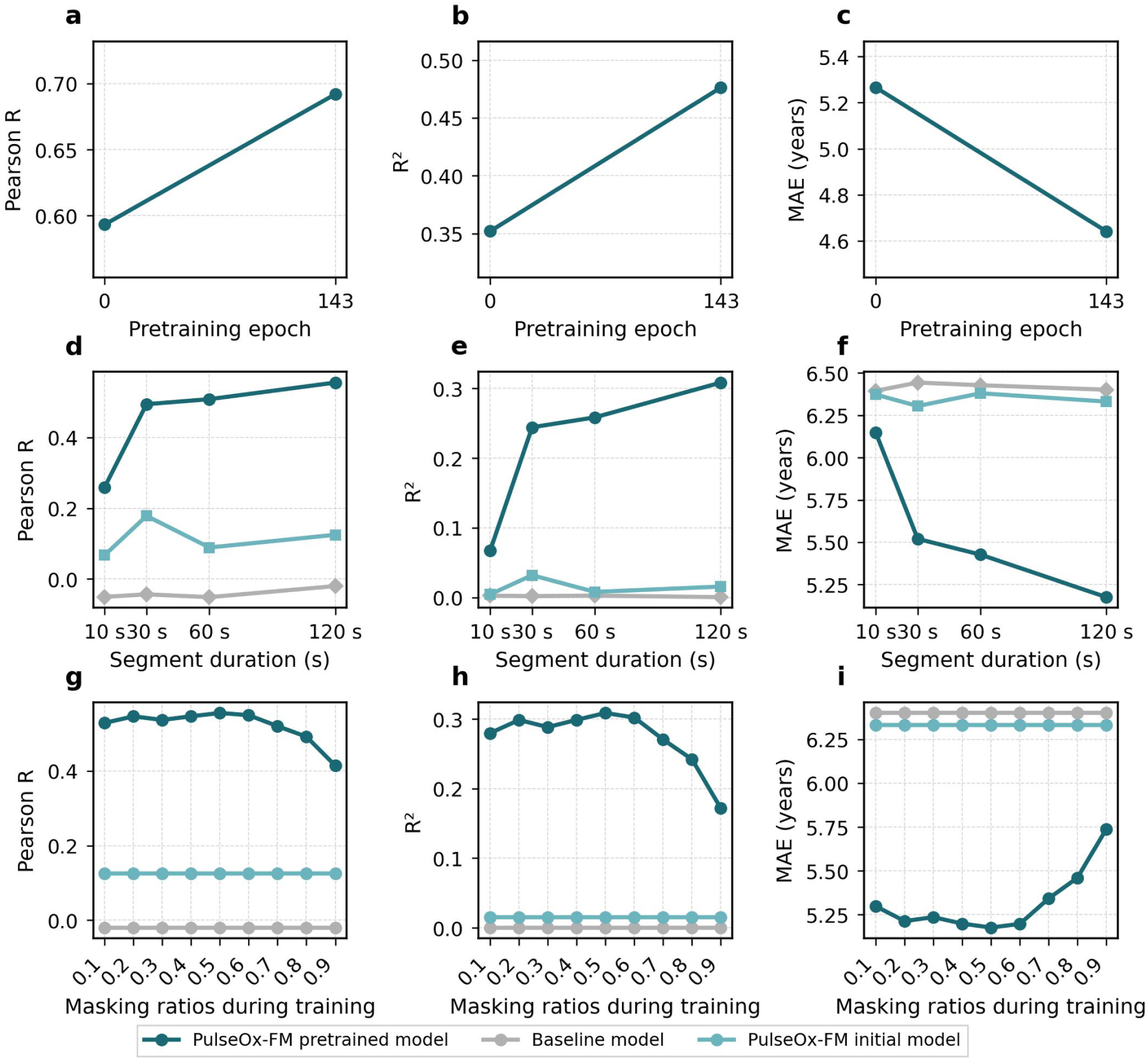
Impact of model training parameters and input configuration. **a–c**, PulseOx-FM performance metrics (Pearson *r*, R^2^ and MAE) for age prediction relative to the number of pretraining epochs. **d–f**, Performance variation across different input segment lengths, ranging from 10 s to 120 s. **g–i**, Evaluation of age prediction performance across different training masking ratios (0.1 to 0.9). Results are compared against a Fourier baseline model (gray) and PulseOx-FM initial model before pretraining (light teal).

**Extended Data Fig. 3.**
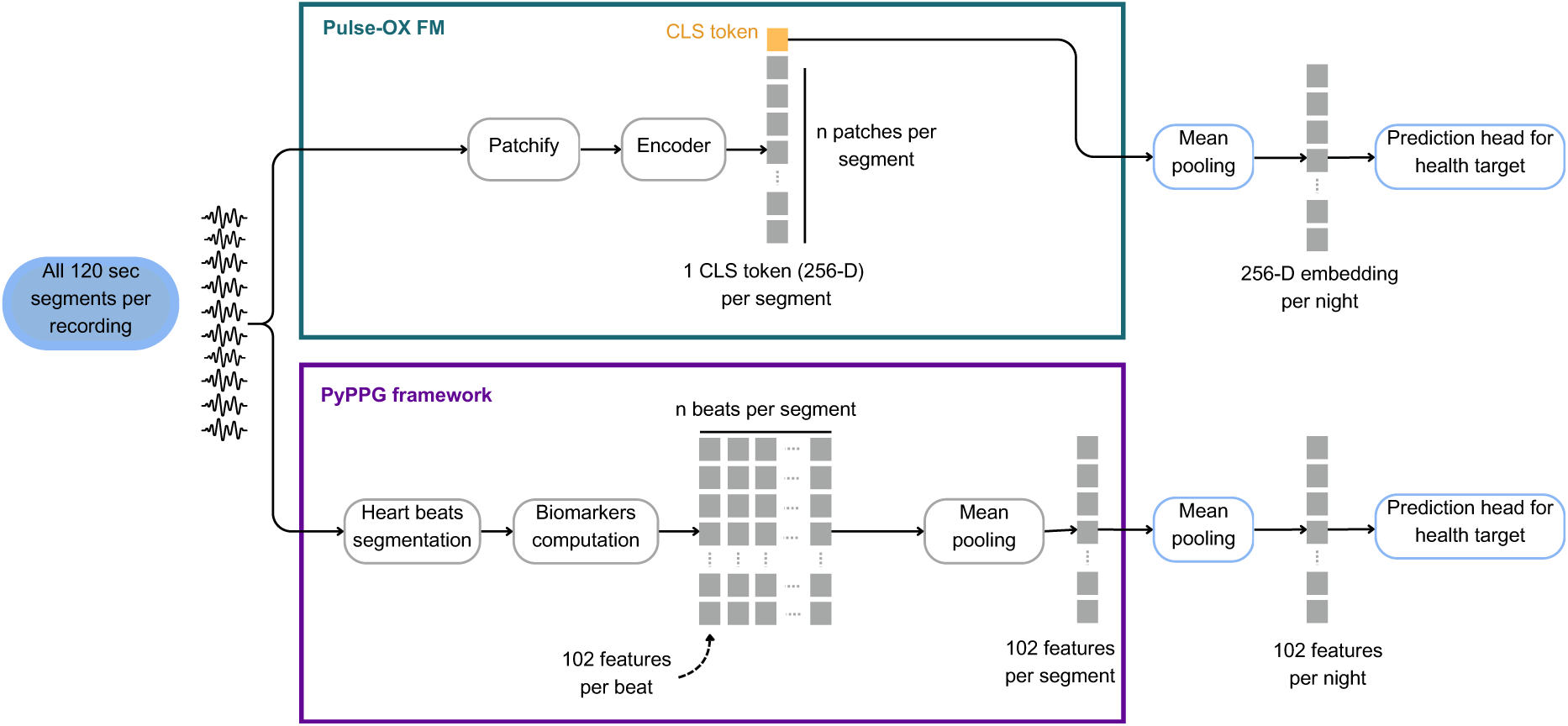
Methodological comparison of PulseOx-FM and PyPPG frameworks for downstream predictions. Schematic representation of the tokenization and pooling strategies used in the foundation model versus the biomarker-based computation in PyPPG.

**Extended Data Fig. 4.**
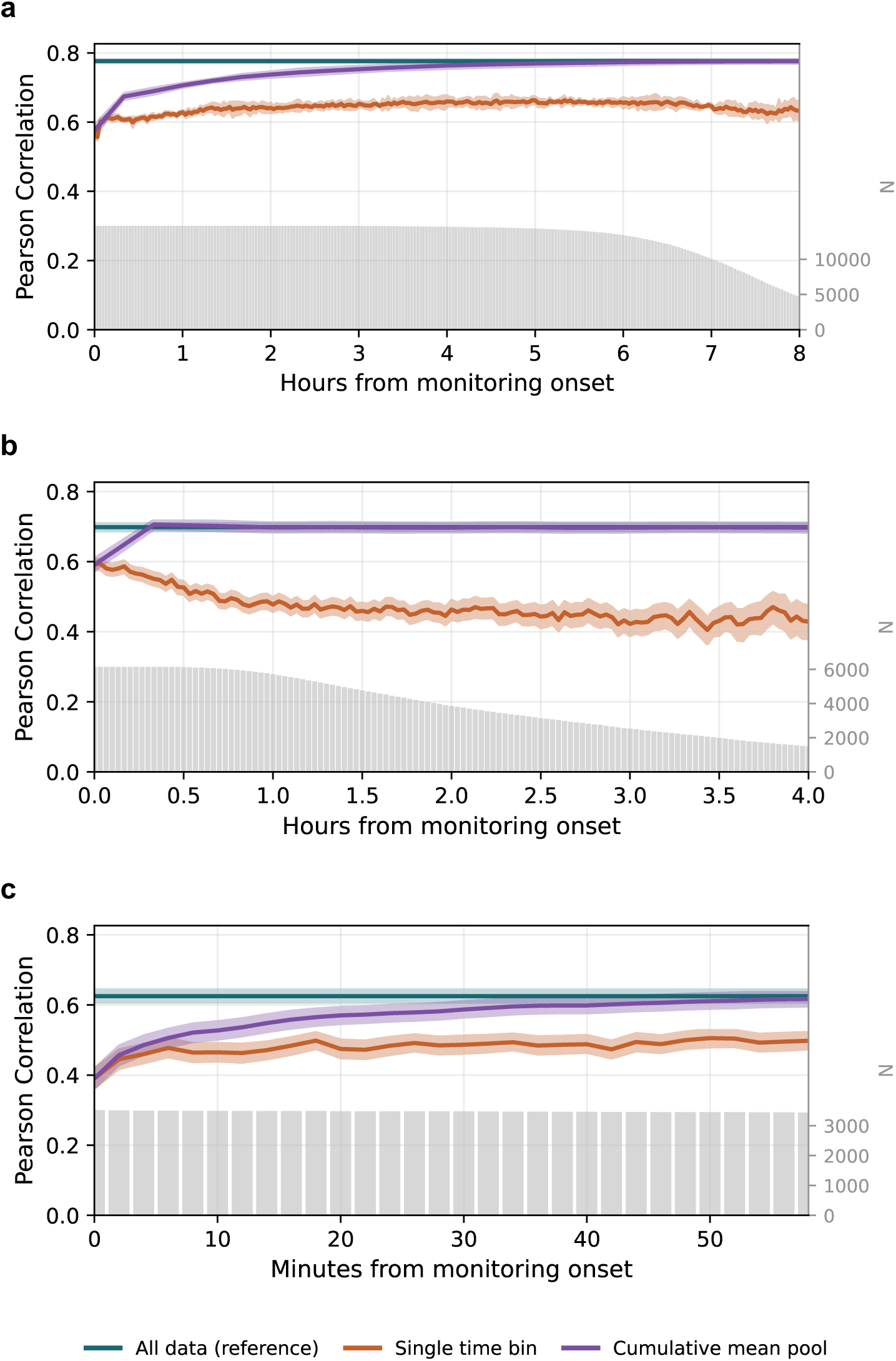
Temporal stability of age-prediction performance across three cohorts. Pearson *r* for age prediction from a single time bin (orange), from embeddings pooled cumulatively from the onset of monitoring (purple) and from all data (reference, teal). Grey bars give the number of participants per bin. **a**, HPP overnight sleep studies against hours from monitoring onset, in 2-minute segments. The single-segment line varies only between *r* = 0.56 and 0.66, and the cumulative pool rises from 0.58 to 0.78 by 8 h. **b**, VitalDB surgical cases against hours from the start of intraoperative monitoring. The cumulative pool reaches 0.71 after 20 minutes. **c**, MIMIC-III intensive-care admissions against minutes from the onset of monitoring, in 2-minute segments. The cumulative pool rises from 0.39 at the first segment to 0.62 by 58 minutes, reaching the all-data reference of 0.63. Bands are 95% cluster-bootstrap intervals from 2,000 resamples.

**Extended Data Fig. 5.**
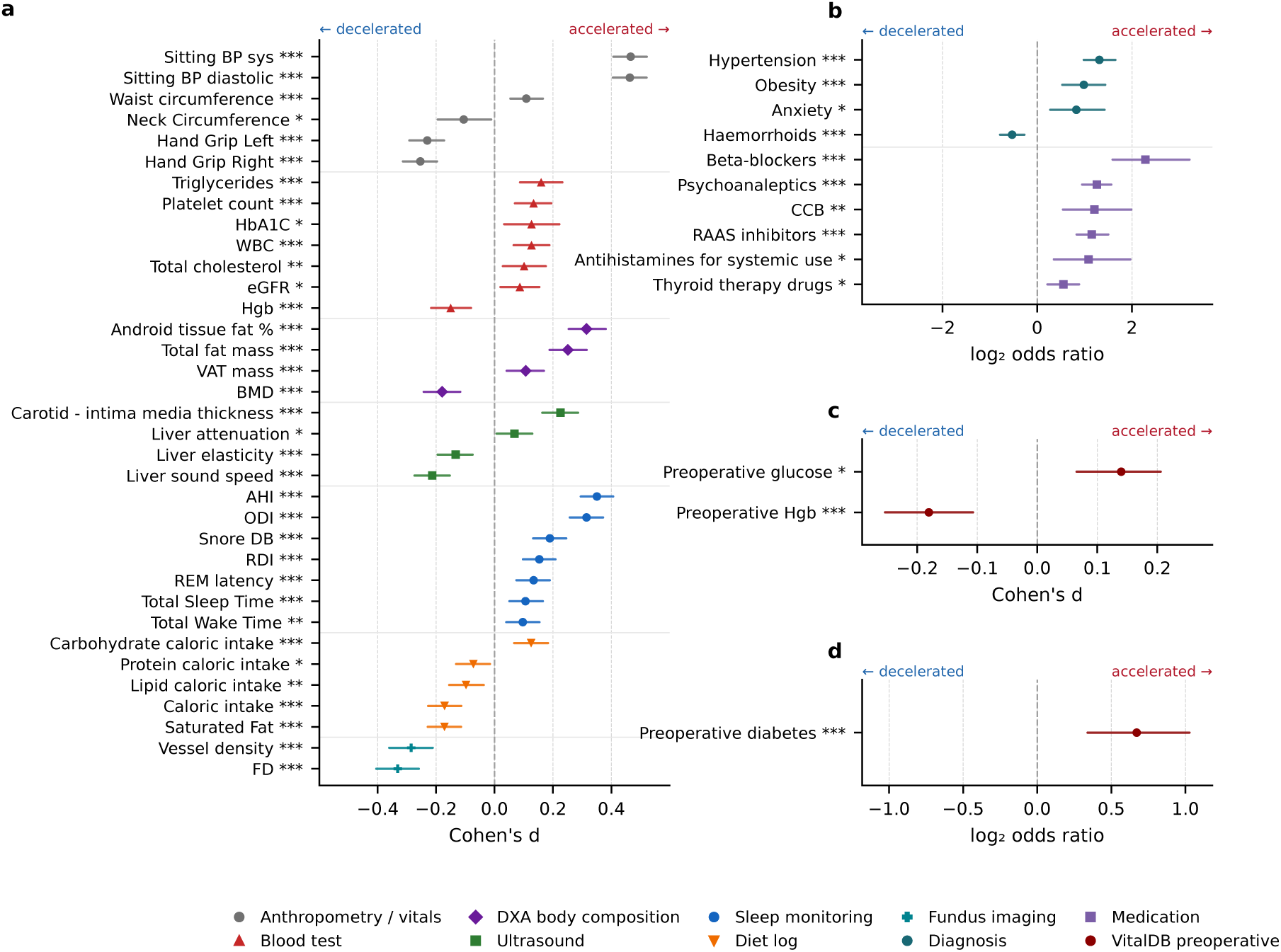
Phenotypes of accelerated versus decelerated agers. **a, b**, HPP sleep cohort: continuous measures (**a**) and diagnoses and medications (**b**) (n = 9,426 participants with an age label). **c, d**, VitalDB surgical cohort: continuous (**c**) and binary (**d**) targets (n = 6,153 cases). Groups are the top (accelerated) and bottom (decelerated) quartiles of the agegap residual (predicted age regressed on chronological age). Markers show the accelerated-minus-decelerated effect size, Cohen’s *d* for continuous measures and log_2_ odds ratio for binary targets, and whiskers are 95% bootstrap confidence intervals (2,000 resamples). Markers right of the dashed zero line are higher in accelerated agers. Only targets significant after Benjamini–Hochberg FDR correction within each task family are shown: 45 of 107 tested targets in HPP and 3 of 5 in VitalDB. Tests are the two-sided Mann–Whitney *U* test (continuous) and the two-sided Fisher exact test (binary); ∗*q* < 0.05, ∗∗*q* < 0.01, ∗∗∗*q* < 0.001. Abbreviations: BMD, bone mineral density; CCB, calcium channel blockers; FD, fractal dimension; HbA1C, haemoglobin A1c; RAAS, renin–angiotensin–aldosterone system.

**Extended Data Fig. 6.**
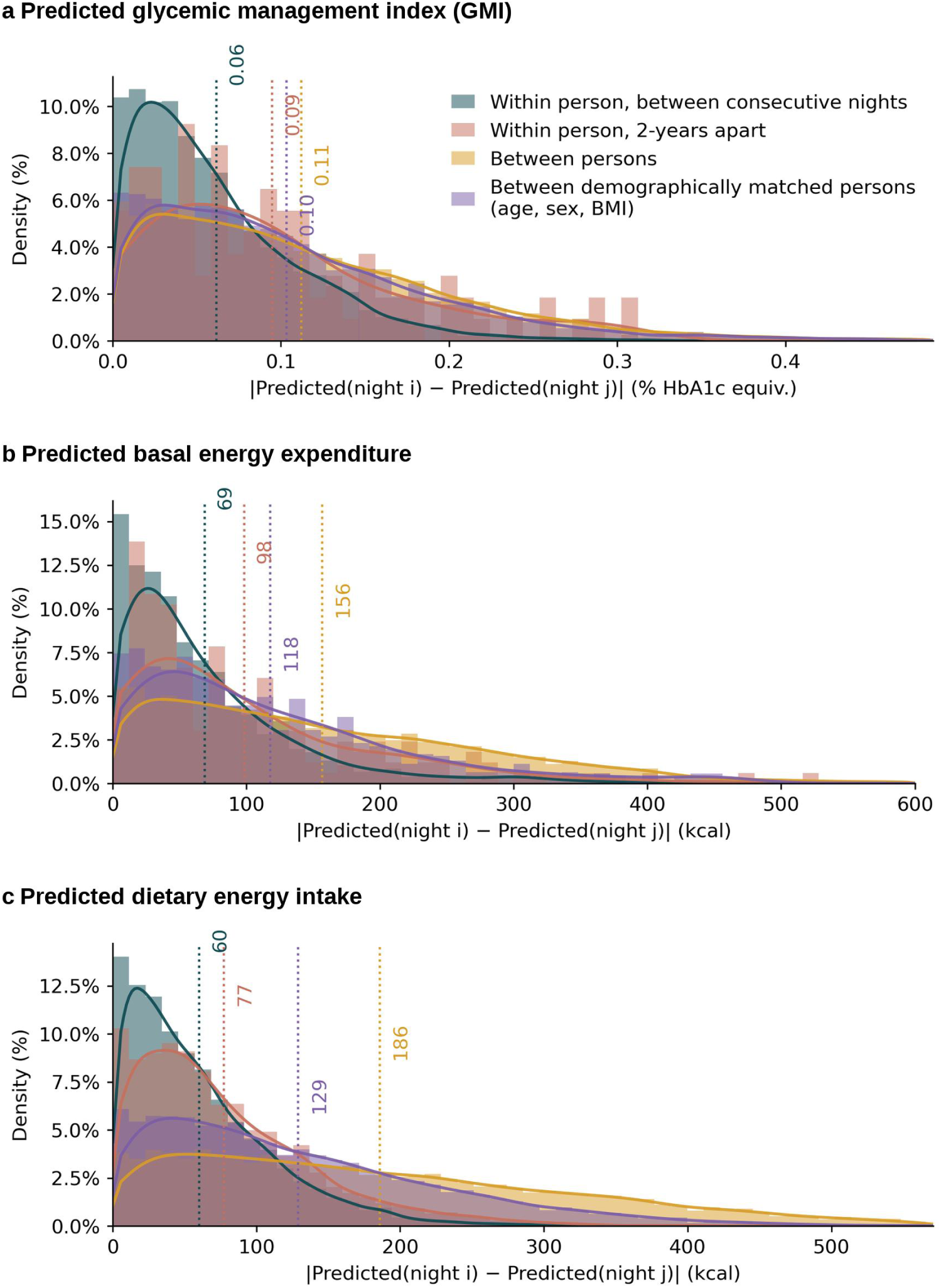
Within- and between-person variability of next-day metabolic predictions. **a–c,** Distributions of the absolute difference between PulseOx-FM predicted values for pairs of sleep recordings, stratified by participant relationship: within the same participant on consecutive nights (dark teal), within the same participant approximately two years apart (coral), between different unmatched participants (gold), and between demographically matched participants (age, sex, and BMI; purple). Panels show prediction-difference distributions for glycemic management index (a), basal energy expenditure (b), and dietary energy intake (c). Histograms represent density; solid curves show kernel density estimates; dotted vertical lines indicate group means. Within-person prediction differences are markedly smaller than between-person differences, including those between demographically matched individuals, that PulseOx-FM sleep embeddings encode individual-specific metabolic signatures not explained by age, sex, or BMI alone.

## Supplementary Material

**Supplementary Figure 1.**
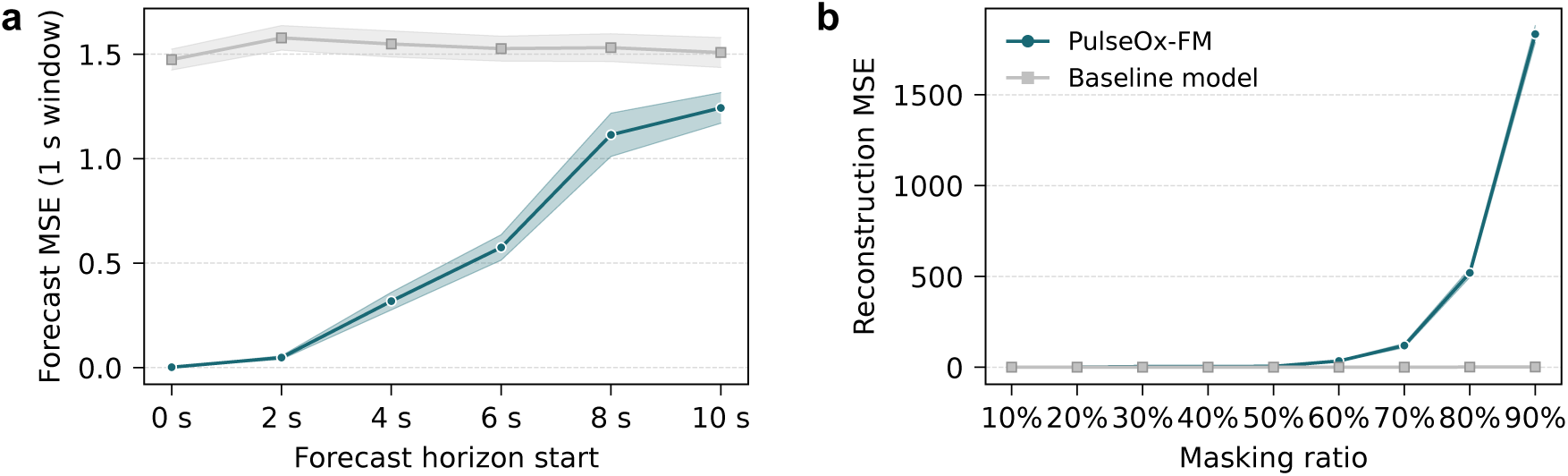
Error analysis of physiological signal reconstruction. **a,** Forecast mean squared error (MSE) evaluated over a 1-second window for PulseOx-FM (teal) and a Fourier-based baseline model (gray) across increasing forecast horizons from 0 to 10 seconds. **b**, Reconstruction MSE relative to the masking ratio (10% to 90%) for PulseOx-FM and the baseline model. Data is presented as mean values across the 1000 evaluated samples from the held-out test set, with shaded regions representing the 95% confidence intervals. MSE, mean squared error.

**Supplementary Figure 2.**
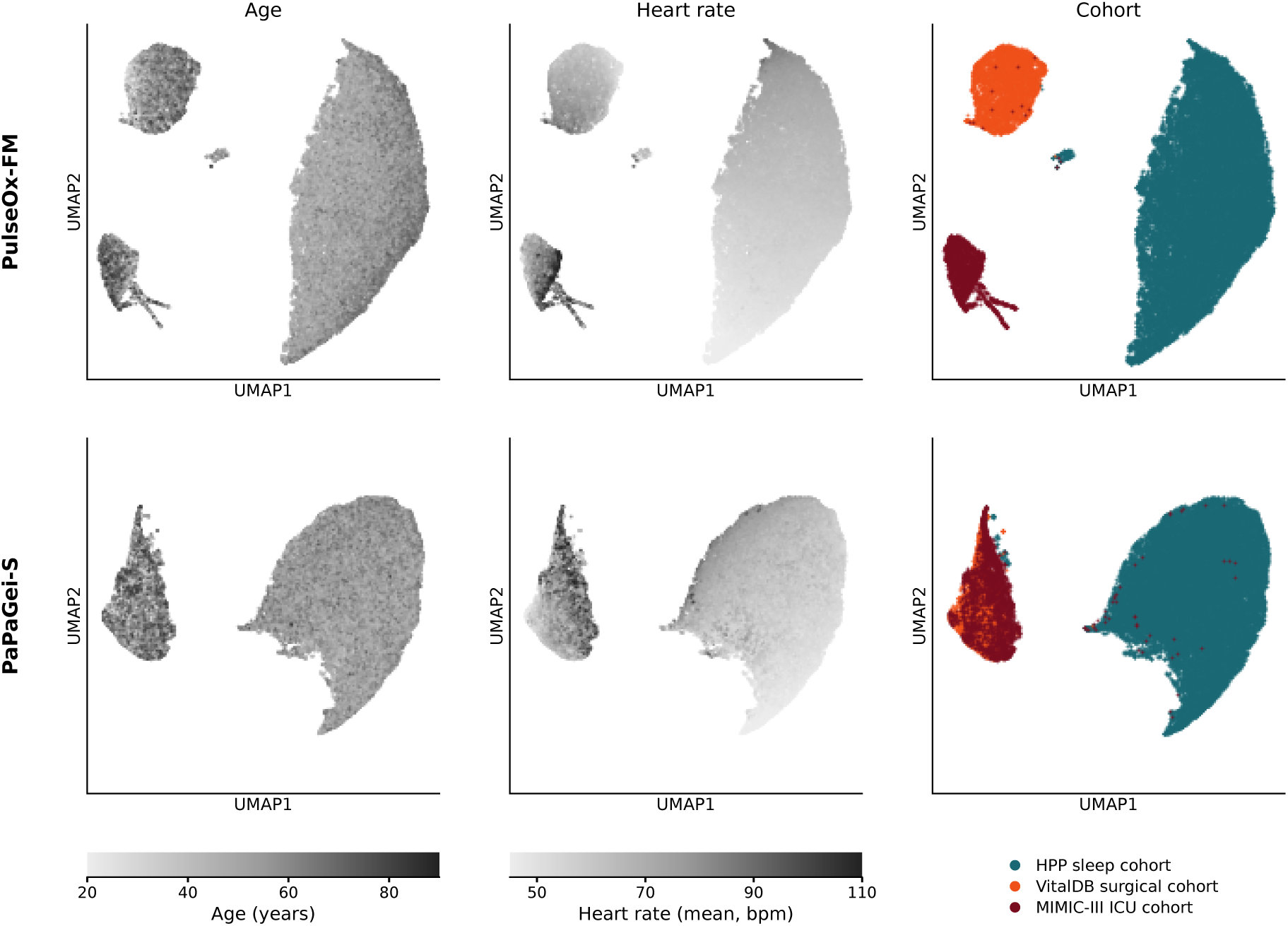
UMAP structure of two learned representations across three cohorts. UMAP projections of PulseOx-FM embeddings (256 dimensions; top) and PaPaGei-S embeddings (512 dimensions; bottom), each fitted jointly on the HPP sleep, VitalDB surgical and MIMIC-III intensive-care cohorts. The columns show the same projection coloured by chronological age (left), by mean heart rate (middle) and by cohort (right: teal HPP, orange VitalDB, dark red MIMIC-III). In the left and middle columns, every cohort shares one grey scale. UMAP preserves local neighbourhoods rather than interpretable axes: the separation of the cohorts shows the cohort shift, and the gradients within each region show retained physiological structure.

**Supplementary Figure 3.**
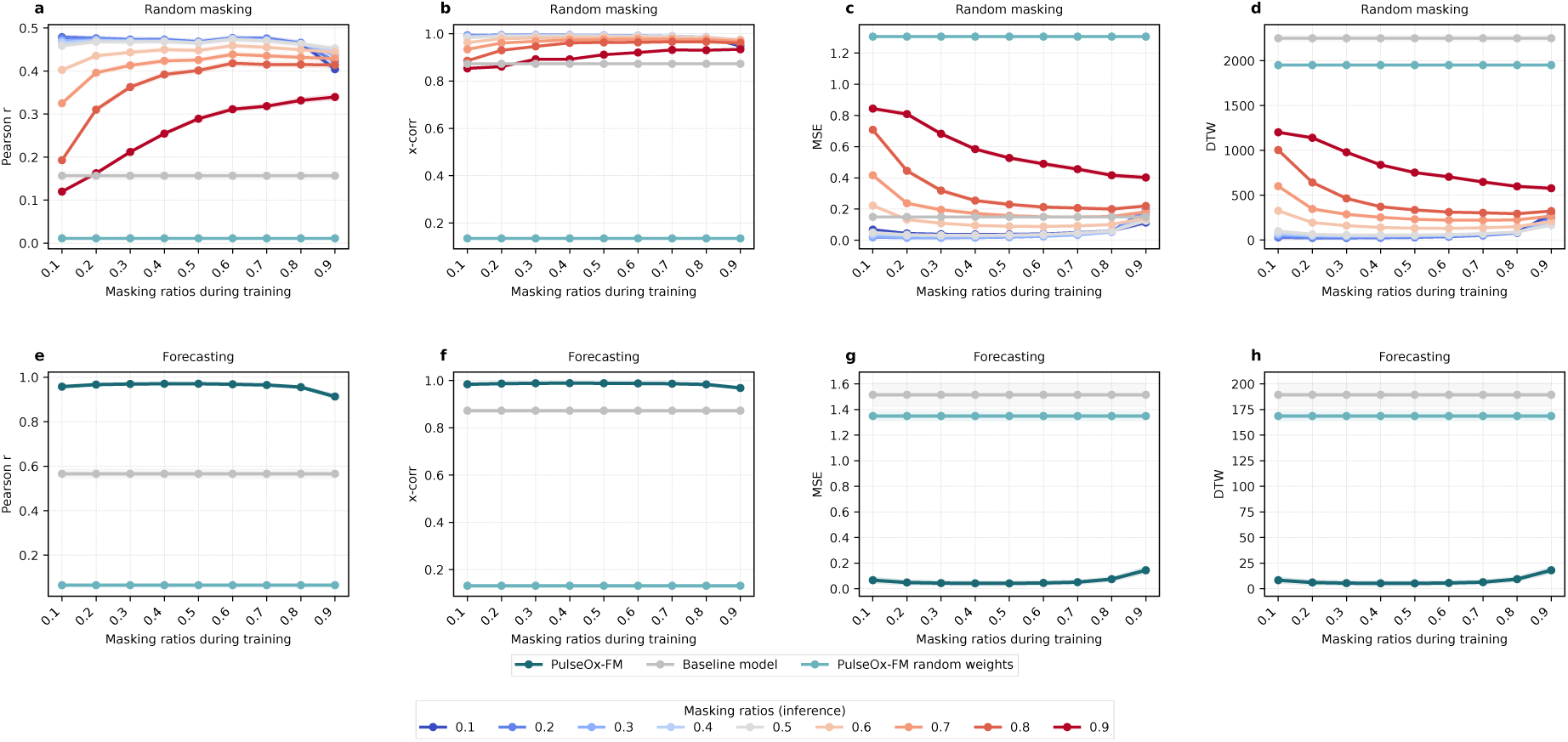
Comprehensive evaluation of model performance across varying masking and training configurations. **a–d,** Quantitative assessment of random masking performance using Pearson r (a), cross-correlation (b), mean squared error (c), and dynamic time warping (d) across different masking ratios used during training (x-axis) and inference (colored lines). **e–h**, Corresponding performance metrics for signal forecasting tasks. Results compare the optimized PulseOx-FM (teal) against a Fourier-based baseline model (gray) and a PulseOx-FM variant with random weights (light teal). DTW, dynamic time warping; MSE, mean squared error; x-corr, maximum cross-correlation.

**Supplementary Figure 4.**
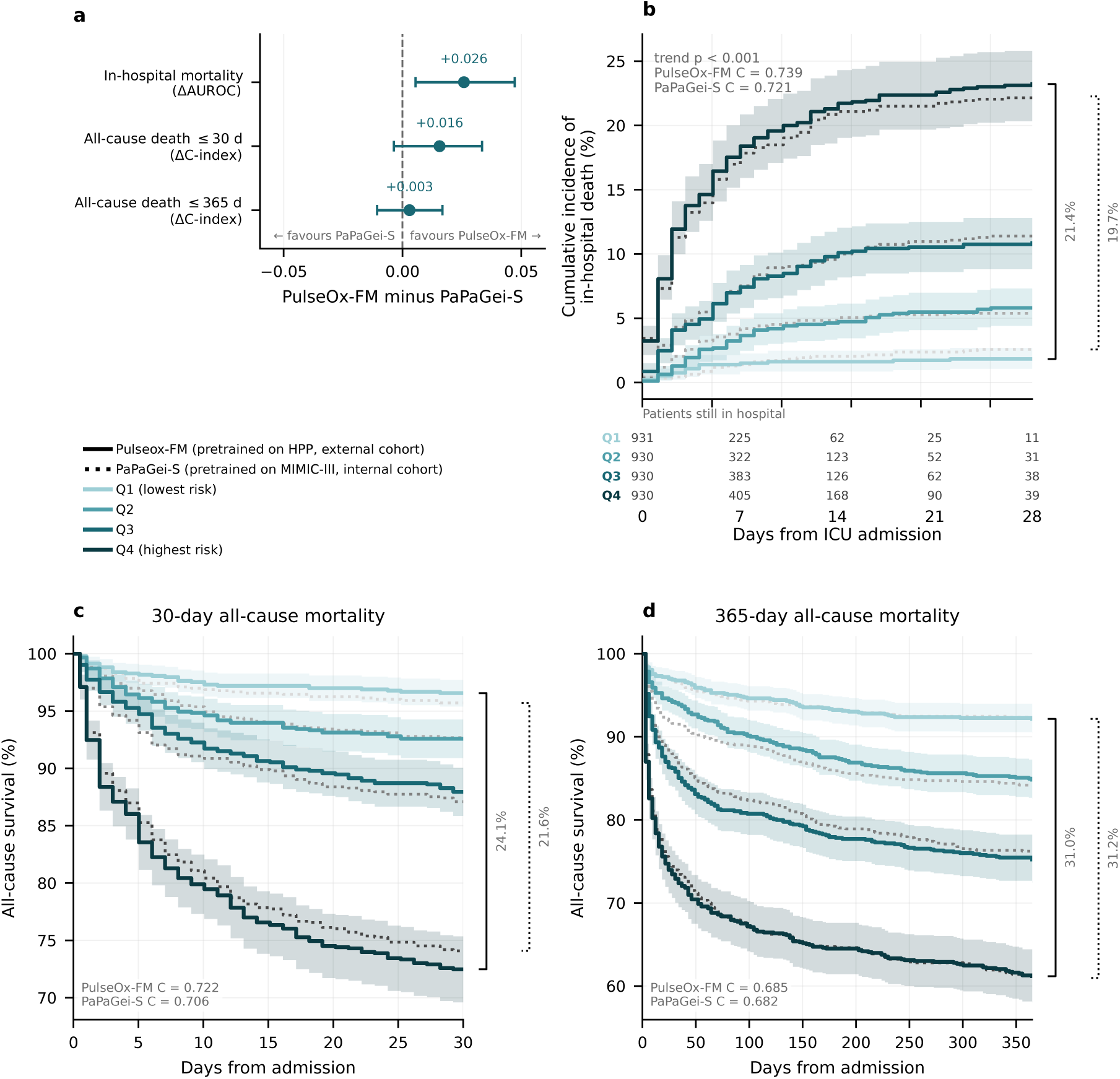
Mortality risk in MIMIC-III, benchmarked against PaPaGei-S. **a**, PulseOx-FM minus PaPaGei-S on identical patients, identical 120-second windows, identical patient-grouped folds and identical prediction heads, with 95% patient-level cluster bootstrap confidence intervals. PaPaGei-S was pretrained on MIMIC-III, whereas PulseOx-FM is applied without fine-tuning. **b**, Cumulative incidence of in-hospital death by quartile of predicted risk (n = 3,721 patients; 402 in-hospital deaths; about 930 per quartile), estimated by the Aalen–Johansen estimator with discharge alive as a competing event, for PulseOx-FM (solid) and PaPaGei-S (dotted). Shaded bands (PulseOx-FM only) are 95% intervals from 1,000 patient-level bootstrap resamples, and the table gives the number of patients still in hospital. Inset: log-rank test for trend and C-index of each encoder. **c, d**, All-cause survival by quartile of predicted risk from embeddings alone at 30 days (**c**) and 365 days (**d**), PulseOx-FM (solid, with 95% Greenwood bands) against PaPaGei-S (dotted). Brackets give the Q1-minus-Q4 separation.

**Supplementary Figure 5.**
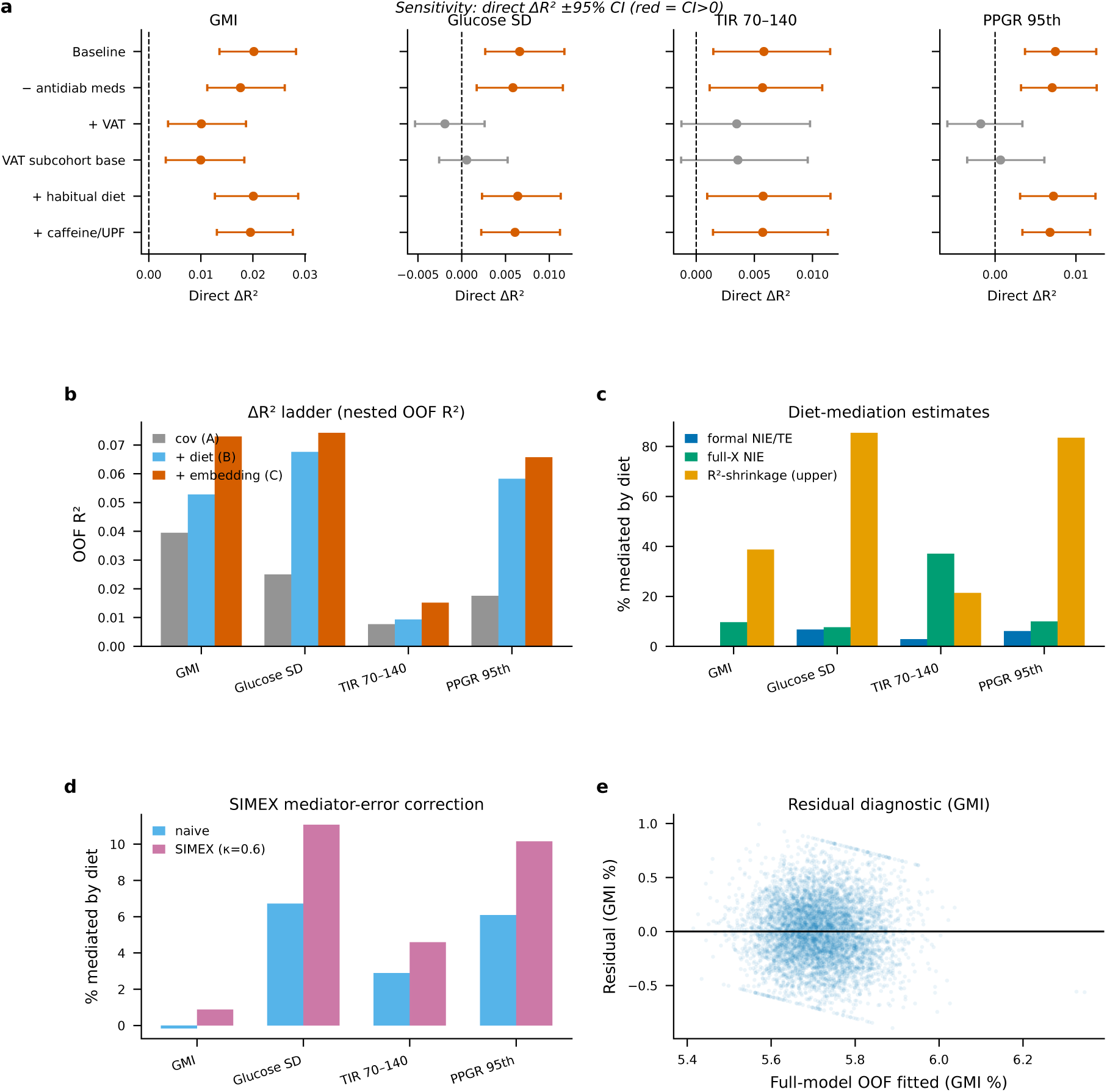
The diet-independent next-day glycemic path is robust to confounder adjustment. **a**, Sensitivity analysis of the direct embedding effect (`Δ`R^2^ of the PulseOx-FM embedding added to demographic covariates plus next-day macronutrient intake) for the four CGM-derived targets (GMI, glucose SD, TIR 70–140 mg/dL, PPGR 95th percentile) under six model specifications: baseline (n = 6,476 participants / 14,526 person-days); excluding anti-diabetic-medication users; adding a visceral-adiposity proxy (android fat, VAT); the same DXA subcohort (n = 4,057) without the visceral-adiposity proxy (selection control); adding participant-mean (habitual) dietary intake; and adding caffeine and ultra-processed-food mediators. Points show out-of-fold `Δ`R^2^; horizontal bars show 95% confidence intervals from 1,000 subject-level bootstrap resamples; red denotes a confidence interval excluding zero, grey one including zero. The matched no-VAT control isolates subcohort selection from VAT confounding. **b**, Nested out-of-fold R^2^ ladder per target: demographic covariates only (grey), plus next-day macronutrients (light blue), plus PulseOx-FM embedding (orange). **c**, Three estimates of the diet-mediated fraction per target: the formal product-of-coefficients natural indirect effect over total effect (NIE/TE, a lower bound because the score is outcome-optimised), the full-embedding NIE using the embedding’s predicted macronutrients, and the R^2^-shrinkage upper bound (1 - direct/total). **d**, SIMEX correction for self-reported-diet measurement error (assumed reliability *κ* = 0.6): diet-mediated fraction before and after correction; the correction raises the mediated fraction modestly while the direct effect is essentially unchanged. **e**, Residual-versus-fitted plot for the full GMI model (out-of-fold), showing homoscedastic residuals. CGM, continuous glucose monitoring; DXA, dual-energy X-ray absorptiometry; `Δ`R^2^, change in coefficient of determination; GMI, glucose management indicator; NIE, natural indirect effect; PPGR, postprandial glucose response; SD, standard deviation; SIMEX, simulation-extrapolation; TE, total effect; TIR 70–140, time in glucose range 70–140 mg/dL; VAT, visceral adipose tissue.

**Supplementary Figure 6.**
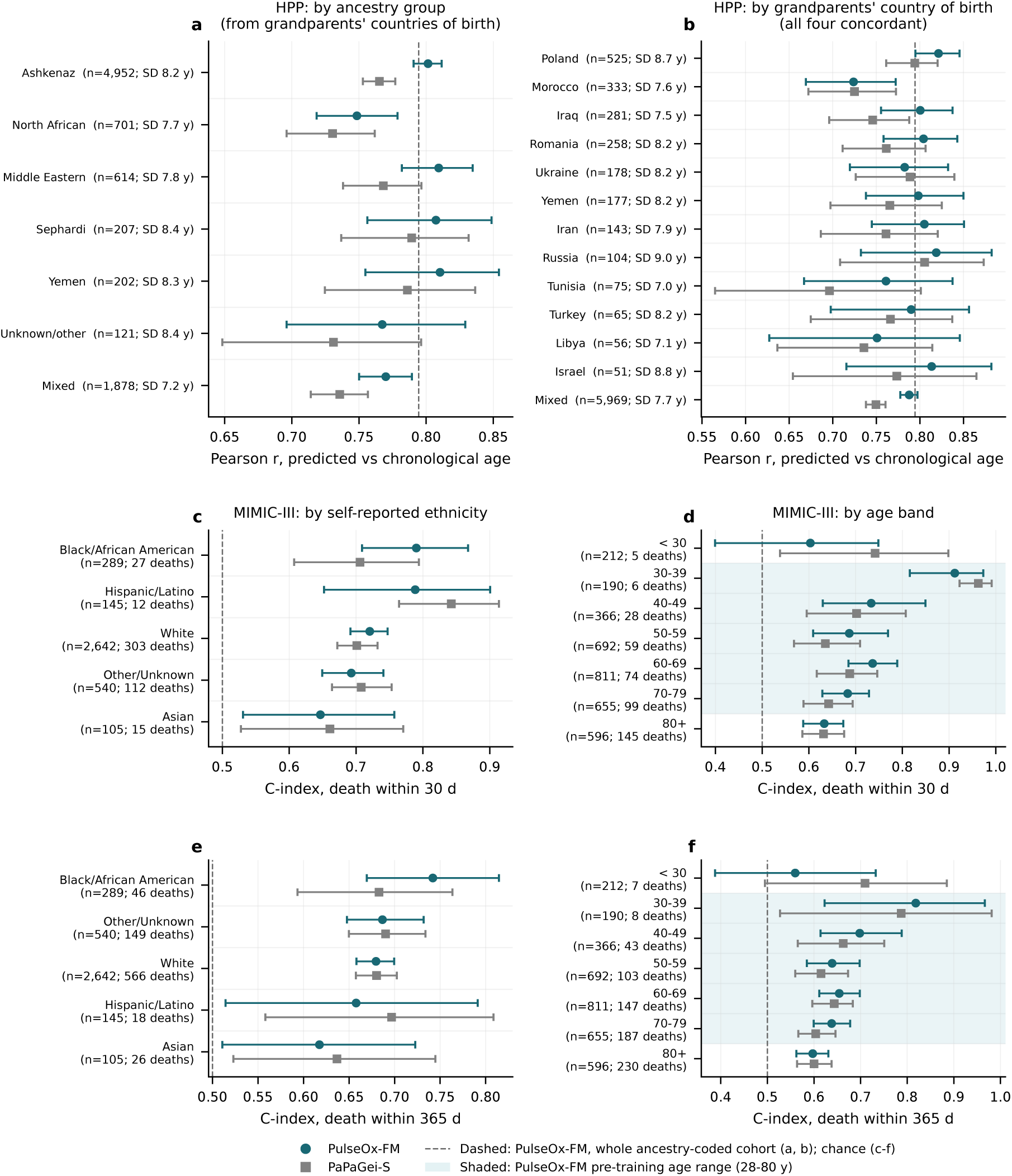
Subgroup analyses of age prediction in HPP and of mortality discrimination in MIMIC-III. **a, b**, Pearson *r* between predicted and chronological age in the HPP sleep cohort, with 95% participant-clustered bootstrap confidence intervals, for PulseOx-FM (circles) and for PaPaGei-S fitted under an identical protocol (squares). Panel **a** is by ancestry group, derived from the four grandparents’ countries of birth; participants whose grandparents’ origins disagree are labelled mixed. Panel **b** is by country, where all four grandparents were born in the same country. The number of participants and the standard deviation of chronological age are given for each group, because a within-group correlation depends on the group’s age spread. The dashed line marks PulseOx-FM in the whole ancestry-coded cohort (*r* = 0.794, n = 8,678). **c–f**, Harrell’s C-index for all-cause death within 30 days (**c**, **d**) and 365 days (**e**, **f**) in the MIMIC-III intensive-care cohort (n = 3,721). Values are for Cox models on PulseOx-FM embeddings applied without fine-tuning (circles) and on PaPaGei-S embeddings (squares), by self-reported ethnicity (**c**, **e**) and by age band (**d**, **f**), with 95% patient-level bootstrap confidence intervals; the dashed line is chance. Shading marks the age range covered by PulseOx-FM pretraining (28–80 years). PaPaGei-S was pretrained on MIMIC-III.

**Supplementary Table 1.** Medication categories used in the medication prediction analyses. Medication targets are listed by ATC-derived category with prevalence among the participants analysed in Fig. 3 (one visit per participant). Prevalence is reported as positive participants/total participants and percentage.

| Medication_Category | Prevalence_N_Total | Prevalence_Percent |
| --- | --- | --- |
| Agents Acting on the Renin-Angiotensin System | 645/10647 | 6.1 |
| Antianemic Preparations | 668/10647 | 6.3 |
| Antigout Preparations | 83/10647 | 0.8 |
| Antihistamines for Systemic Use | 158/10647 | 1.5 |
| Antithrombotic Agents | 340/10647 | 3.2 |
| Calcium Channel Blockers | 151/10647 | 1.4 |
| Drugs for Acid Related Disorders | 382/10647 | 3.6 |
| Drugs for Obstructive Airway Diseases | 157/10647 | 1.5 |
| Drugs for Treatment of Bone Diseases | 78/10647 | 0.7 |
| Drugs Used in Diabetes | 202/10647 | 1.9 |
| Lipid Modifying Agents | 1400/10647 | 13.1 |
| Mineral Supplements | 768/10647 | 7.2 |
| Psychoanaleptics | 888/10647 | 8.3 |
| Psycholeptics | 116/10647 | 1.1 |
| Sex Hormones and Modulators of the Genital System | 533/10647 | 5.0 |
| Thyroid Therapy Drugs | 604/10647 | 5.7 |
| Urologicals | 204/10647 | 1.9 |
| Beta-Adrenergic Blocking Agents | 198/10647 | 1.9 |

**Supplementary Table 2.** Disease categories used in the disease prediction analyses. Disease targets are listed within the prespecified disease categories with prevalence among the participants analysed in Fig. 3 (one visit per participant). Prevalence is reported as positive participants/total participants and percentage.

| Disease_Category | Disease_Used_in_Analysis | Prevalence_N_Total | Prevalence_Percent |
| --- | --- | --- | --- |
| Cardiovascular disorders | Heart valve disease | 74/5669 | 1.3 |
| Cardiovascular disorders | Hypertension | 673/5669 | 11.9 |
| Metabolic, endocrine and reproductive disorders | Fatty Liver Disease | 346/5669 | 6.1 |
| Metabolic, endocrine and reproductive disorders | Gout | 69/5669 | 1.2 |
| Metabolic, endocrine and reproductive disorders | Hyperlipidemia | 1383/5669 | 24.4 |
| Metabolic, endocrine and reproductive disorders | Obesity | 334/5669 | 5.9 |
| Metabolic, endocrine and reproductive disorders | Polycystic Ovary Disease | 142/5669 | 2.5 |
| Metabolic, endocrine and reproductive disorders | Prediabetes | 524/5669 | 9.2 |
| Sleep disorders and mental health | Anxiety | 220/5669 | 3.9 |
| Sleep disorders and mental health | Depression | 202/5669 | 3.6 |
| Sleep disorders and mental health | Insomnia | 98/5669 | 1.7 |
| Sleep disorders and mental health | Sleep Apnea | 181/5669 | 3.2 |
| Gastrointestinal disorders | Anal Fissure | 398/5669 | 7.0 |
| Gastrointestinal disorders | Gallstone Disease | 241/5669 | 4.3 |
| Gastrointestinal disorders | Haemorrhoids | 1288/5669 | 22.7 |
| Gastrointestinal disorders | IBS | 204/5669 | 3.6 |
| Gastrointestinal disorders | Peptic Ulcer Disease | 125/5669 | 2.2 |
| Hematologic and nutritional disorders | Anemia | 276/5669 | 4.9 |
| Hematologic and nutritional disorders | B12 Deficiency | 310/5669 | 5.5 |
| Hematologic and nutritional disorders | G6PD | 66/5669 | 1.2 |
| Neurological and pain conditions | ADHD | 1056/5669 | 18.6 |
| Neurological and pain conditions | Back Pain | 1204/5669 | 21.2 |
| Neurological and pain conditions | Fibromyalgia | 69/5669 | 1.2 |
| Neurological and pain conditions | Headache | 139/5669 | 2.5 |
| Neurological and pain conditions | Migraine | 351/5669 | 6.2 |
| Immune and allergic diseases | Allergy | 1197/5669 | 21.1 |
| Immune and allergic diseases | Asthma | 354/5669 | 6.2 |
| Immune and allergic diseases | Atopic Dermatitis | 195/5669 | 3.4 |
| Immune and allergic diseases | Psoriasis | 167/5669 | 2.9 |
| Other | Fractures | 556/5669 | 9.8 |
| Other | Hearing loss | 222/5669 | 3.9 |
| Other | Oral apthae | 246/5669 | 4.3 |
| Other | Osteoarthritis | 171/5669 | 3.0 |
| Other | Sinusitis | 194/5669 | 3.4 |
| Other | Urinary Tract Stones | 191/5669 | 3.4 |
| Other | Urinary tract infection | 575/5669 | 10.1 |

**Supplementary Table 3.** Age prediction performance and agreement across feature sets and four cohorts. Pearson *r*, R^2^ and mean absolute error (MAE) are shown as mean ± s.d. across three cross-validation seeds. Within each cohort, all feature sets are evaluated on the same rows; N is the number of participants evaluated. Units are participant-visits (HPP), cases (VitalDB) and patients (MIMICIII). Bland–Altman statistics are computed on the seed-averaged predictions, with differences taken against chronological age (Krouwer orientation): bias (predicted minus true age, years, with 95% CI), 95% limits of agreement (LoA; bias ± 1.96 s.d.), their width, and the percentage of participants within them. PaPaGei-S was pretrained on VitalDB and MIMIC-III.

| Dataset | Model or features | N | Pearson $r$ | $R^2$ | MAE (years) | Bias (years) | Bias 95% CI | LoA lower | LoA upper | LoA width | Within LoA (%) |
| --- | --- | --- | --- | --- | --- | --- | --- | --- | --- | --- | --- |
| HPP pretraining set (train + validation) | PulseOx-FM | 9,443 | 0.79 ± 0.0002 | 0.63 ± 0.0004 | 3.88 ± 0.0027 | -0.01 | -0.09 to 0.07 | -9.60 | 9.58 | 19.18 | 94.5 |
| HPP pretraining set (train + validation) | PaPaGei-S | 9,443 | 0.76 ± 0.0008 | 0.57 ± 0.0011 | 4.16 ± 0.0060 | 0.02 | -0.07 to 0.10 | -10.26 | 10.29 | 20.56 | 94.7 |
| HPP pretraining set (train + validation) | PyPPG | 9,443 | 0.74 ± 0.0004 | 0.55 ± 0.0007 | 4.25 ± 0.0004 | -0.12 | -0.21 to -0.04 | -10.63 | 10.38 | 21.00 | 94.9 |
| HPP pretraining set (train + validation) | WatchPAT | 9,443 | 0.64 ± 0.0009 | 0.40 ± 0.0012 | 4.97 ± 0.0040 | 0.06 | -0.04 to 0.16 | -12.13 | 12.24 | 24.38 | 95.0 |
| HPP pretraining set (train + validation) | Ensemble of PulseOx-FM + PyPPG | 9,443 | 0.80 ± 0.0001 | 0.64 ± 0.0003 | 3.85 ± 0.0033 | -0.07 | -0.15 to 0.01 | -9.55 | 9.42 | 18.97 | 94.8 |
| HPP test set | PulseOx-FM | 606 | 0.73 ± 0.0070 | 0.54 ± 0.0103 | 4.38 ± 0.0441 | 0.01 | -0.32 to 0.34 | -10.58 | 10.60 | 21.18 | 94.8 |
| HPP test set | PaPaGei-S | 606 | 0.73 ± 0.0058 | 0.53 ± 0.0085 | 4.41 ± 0.0253 | 0.08 | -0.25 to 0.42 | -10.63 | 10.80 | 21.42 | 94.9 |
| HPP test set | PyPPG | 606 | 0.69 ± 0.0069 | 0.48 ± 0.0095 | 4.58 ± 0.0553 | -0.01 | -0.36 to 0.34 | -11.21 | 11.20 | 22.41 | 94.5 |
| HPP test set | Ensemble of PulseOx-FM + PyPPG | 606 | 0.75 ± 0.0049 | 0.56 ± 0.0072 | 4.27 ± 0.0361 | 0.00 | -0.32 to 0.32 | -10.35 | 10.35 | 20.70 | 94.7 |
| VitalDB cohort | PulseOx-FM | 6,135 | 0.70 ± 0.0014 | 0.49 ± 0.0019 | 8.29 ± 0.0286 | 0.11 | -0.16 to 0.38 | -20.71 | 20.93 | 41.64 | 95.2 |
| VitalDB cohort | PaPaGei-S | 6,135 | 0.67 ± 0.0016 | 0.45 ± 0.0019 | 8.68 ± 0.0132 | 0.71 | 0.43 to 0.98 | -20.88 | 22.30 | 43.18 | 95.1 |
| VitalDB cohort | PyPPG | 6,135 | 0.71 ± 0.0017 | 0.50 ± 0.0023 | 8.19 ± 0.0164 | 0.14 | -0.12 to 0.40 | -20.34 | 20.63 | 40.97 | 95.0 |
| VitalDB cohort | Ensemble of PulseOx-FM + PyPPG | 6,135 | 0.73 ± 0.0002 | 0.54 ± 0.0002 | 7.94 ± 0.0063 | 0.12 | -0.13 to 0.38 | -19.72 | 19.97 | 39.70 | 95.5 |
| MIMIC-III intensive-care cohort | PulseOx-FM | 3,522 | 0.62 ± 0.0019 | 0.39 ± 0.0023 | 10.63 ± 0.0167 | 0.08 | -0.36 to 0.51 | -25.77 | 25.93 | 51.70 | 95.3 |
| MIMIC-III intensive-care cohort | PaPaGei-S | 3,522 | 0.66 ± 0.0009 | 0.43 ± 0.0011 | 10.32 ± 0.0091 | 0.61 | 0.19 to 1.03 | -24.33 | 25.55 | 49.88 | 95.9 |
| MIMIC-III intensive-care cohort | Random-initialised encoder | 3,522 | 0.51 ± 0.0020 | 0.26 ± 0.0018 | 11.79 ± 0.0121 | 0.05 | -0.43 to 0.53 | -28.47 | 28.57 | 57.03 | 95.4 |

**Supplementary Table 4.**
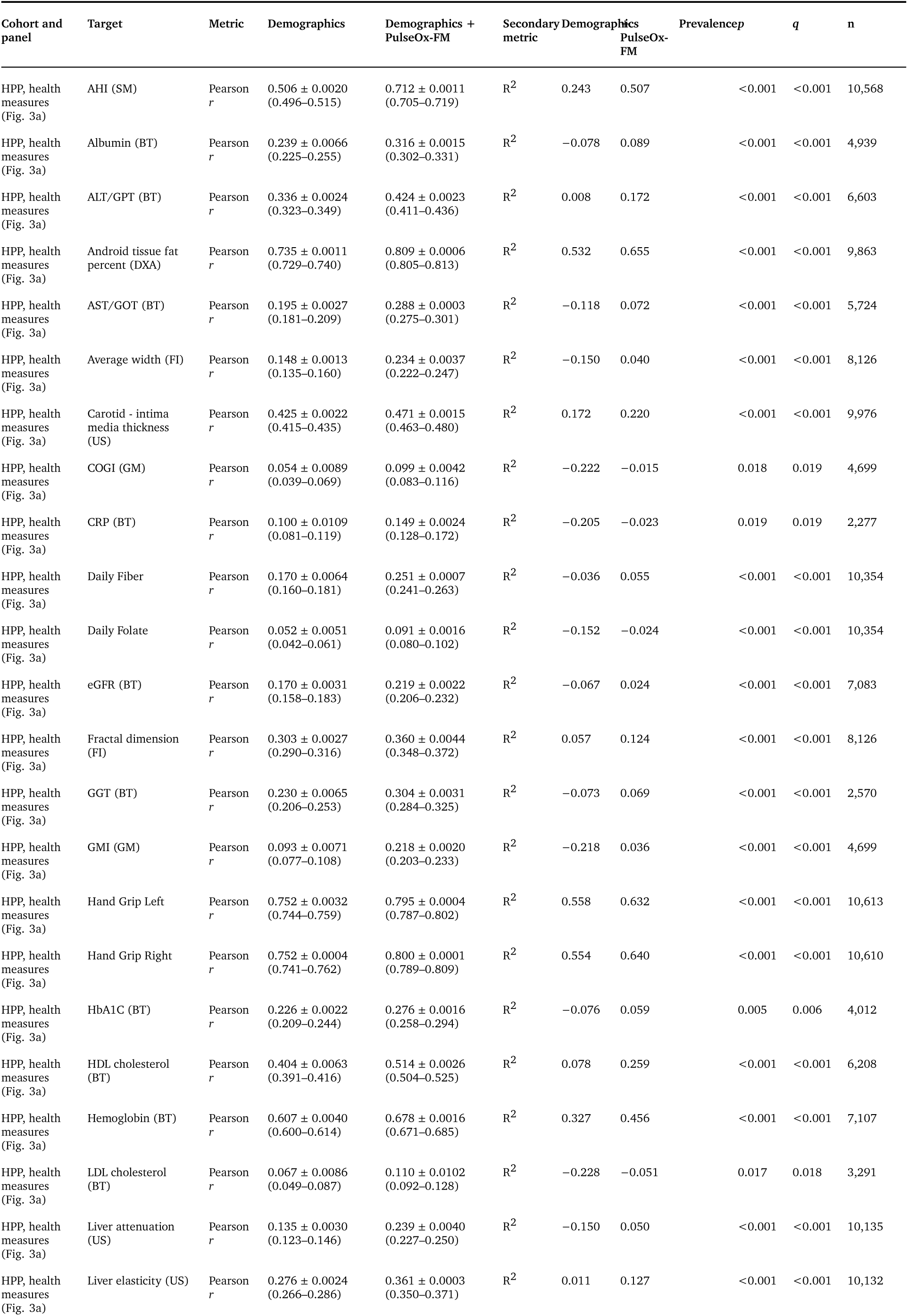
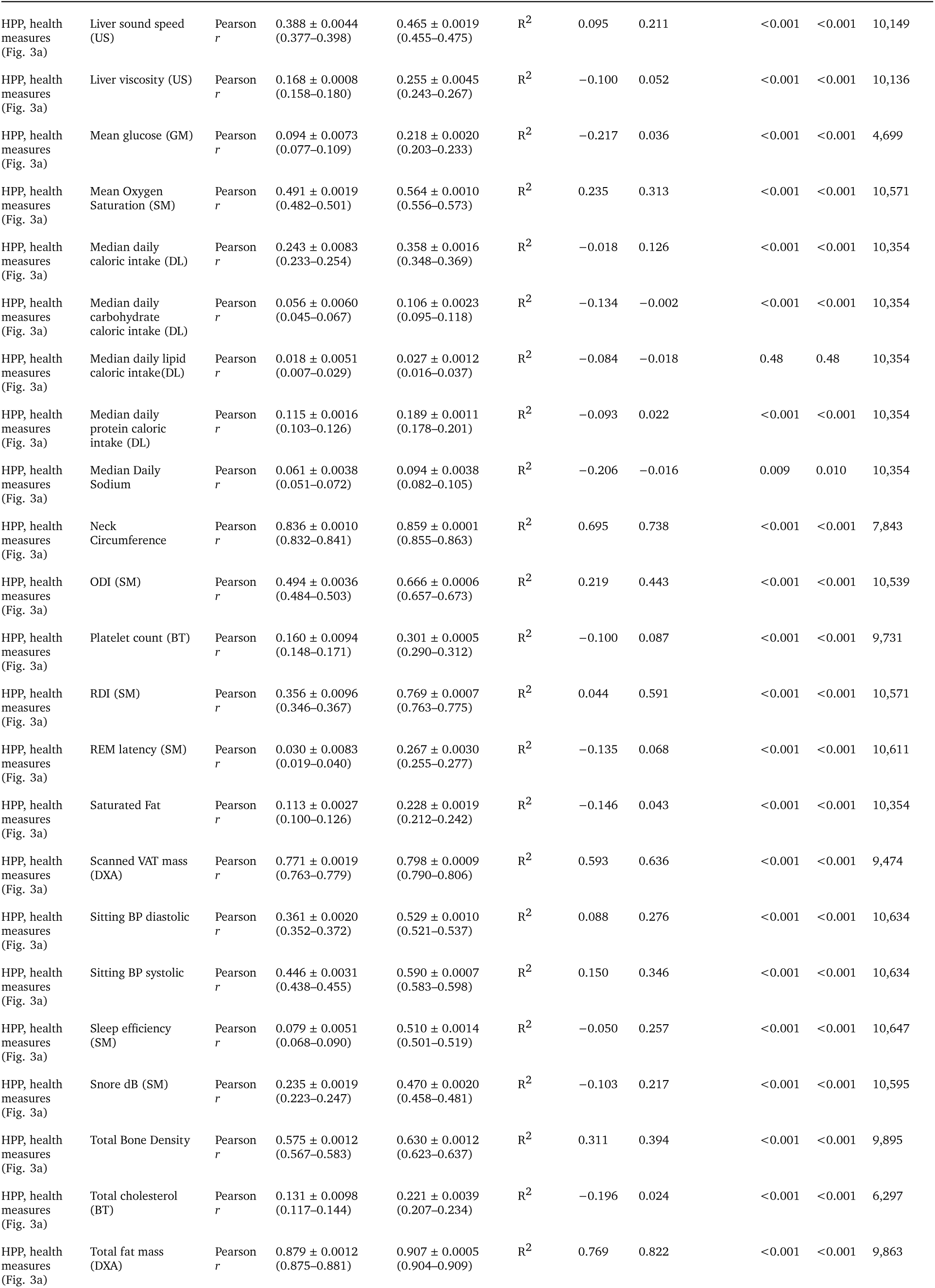
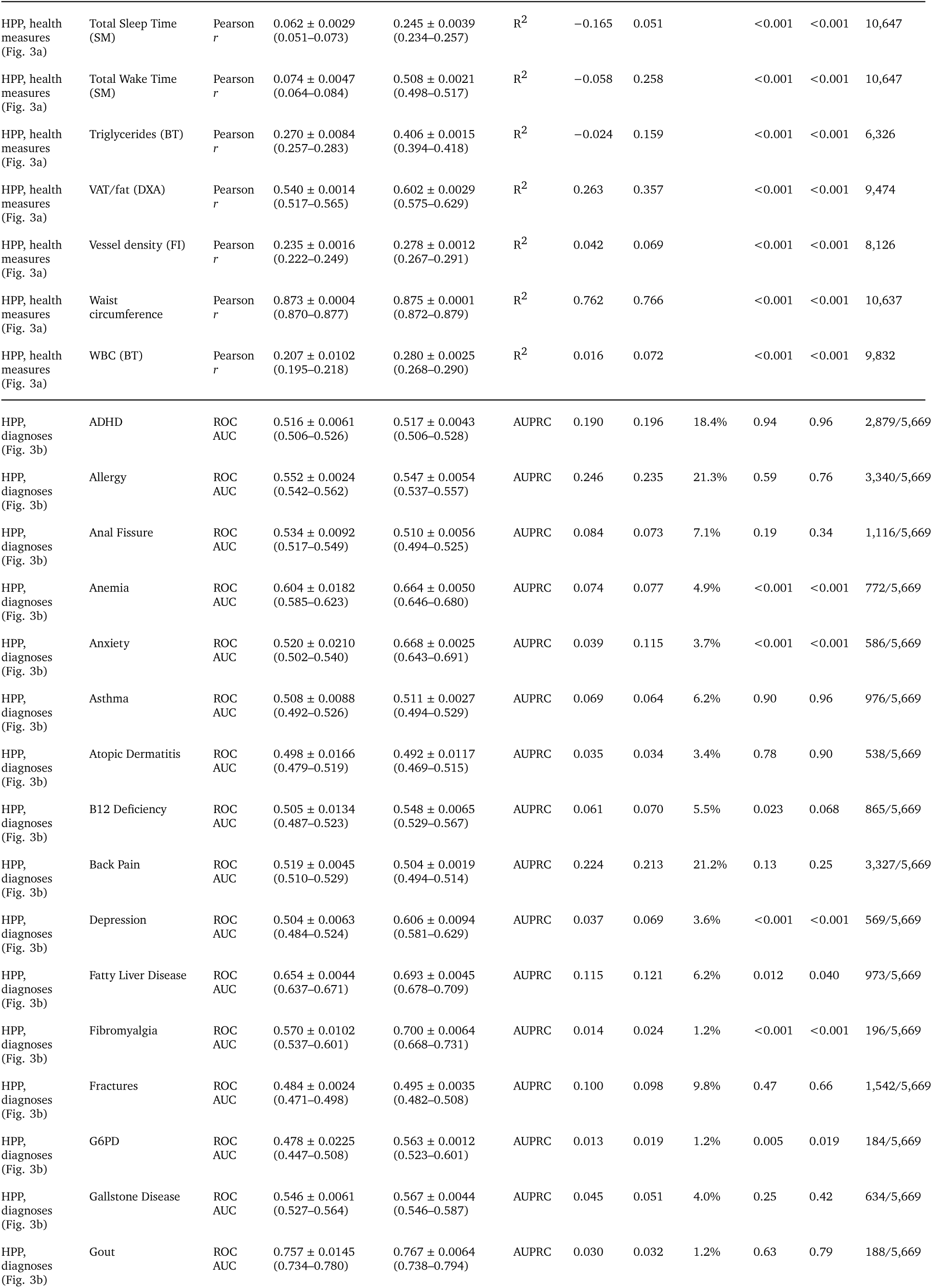
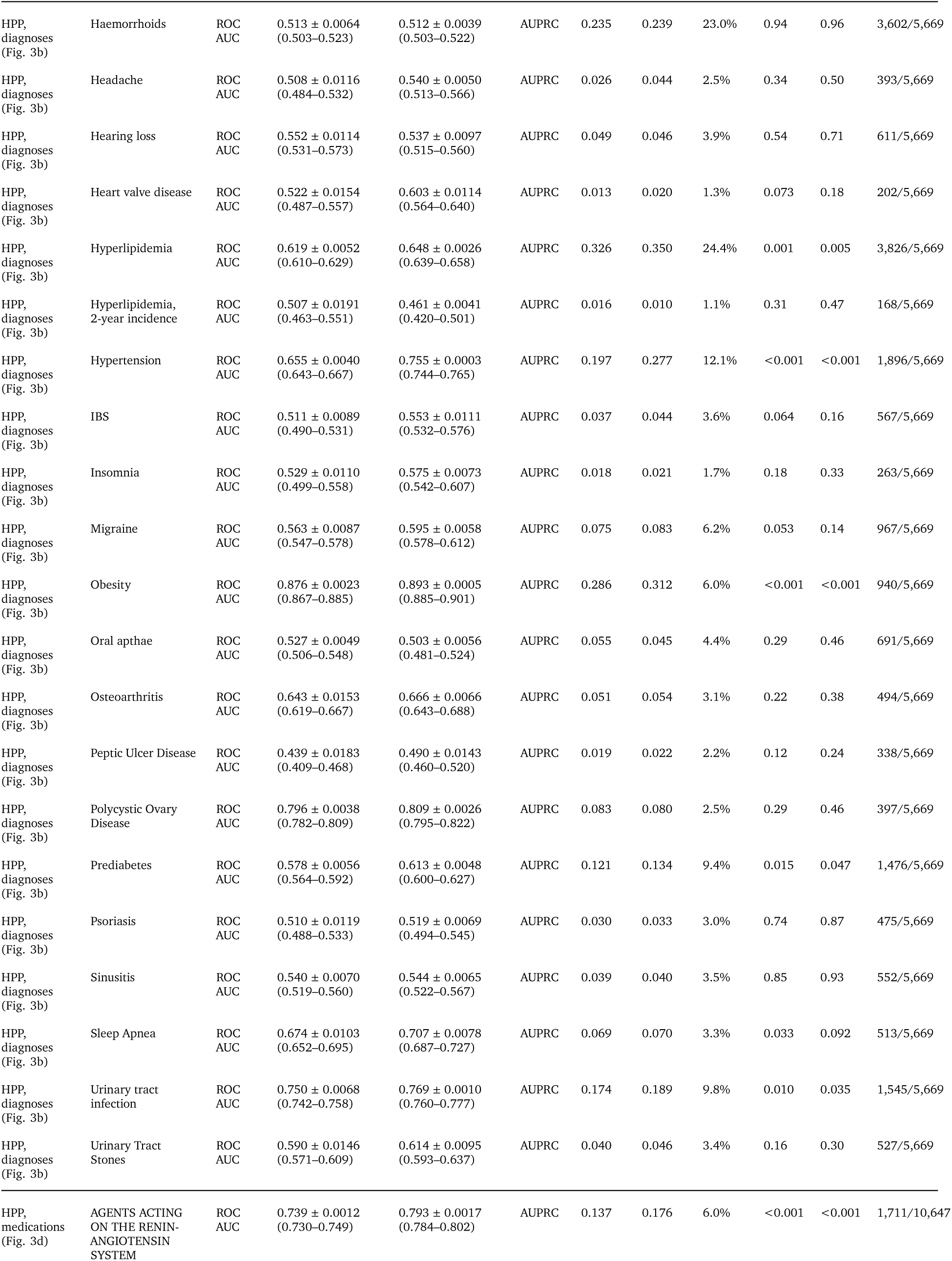
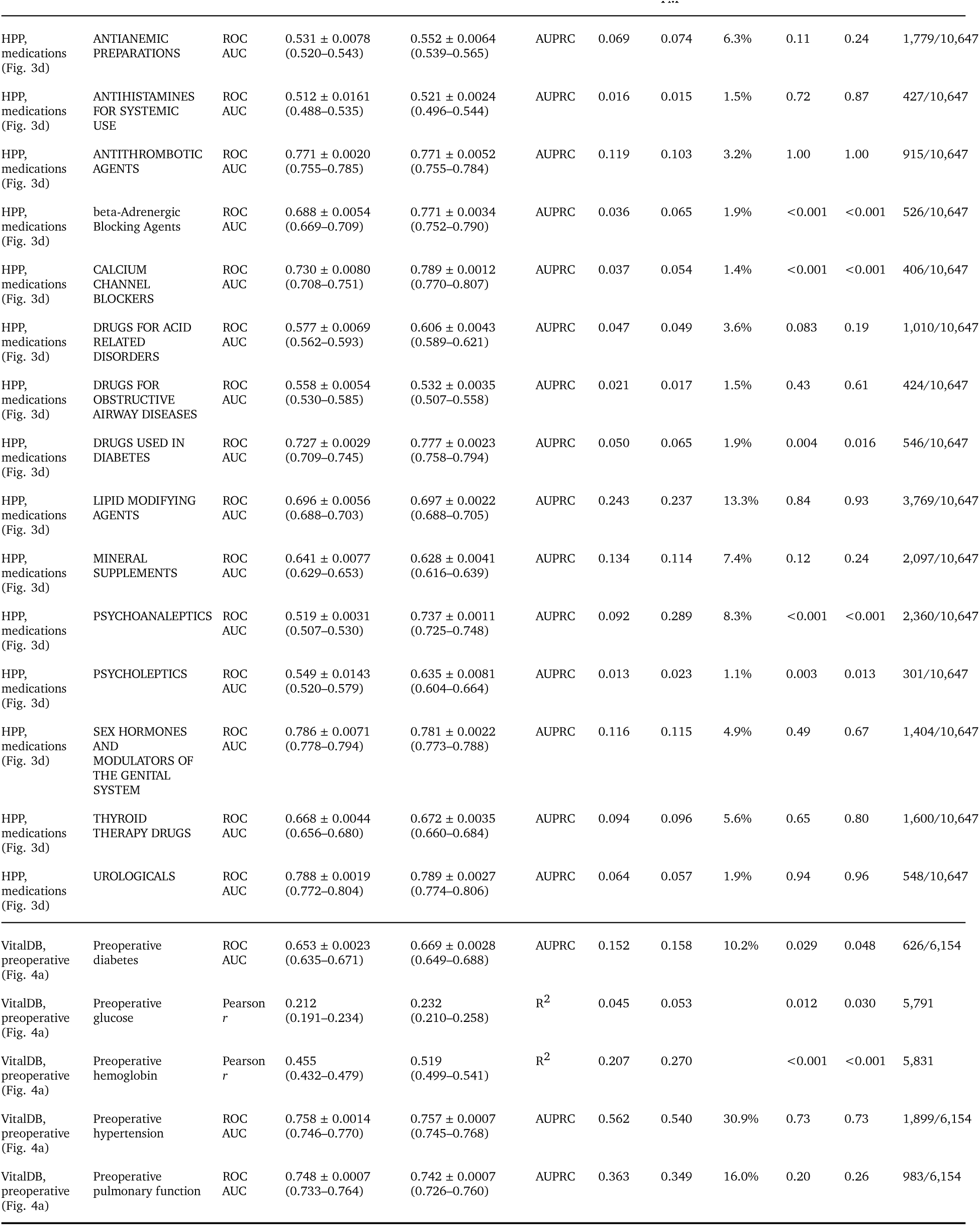
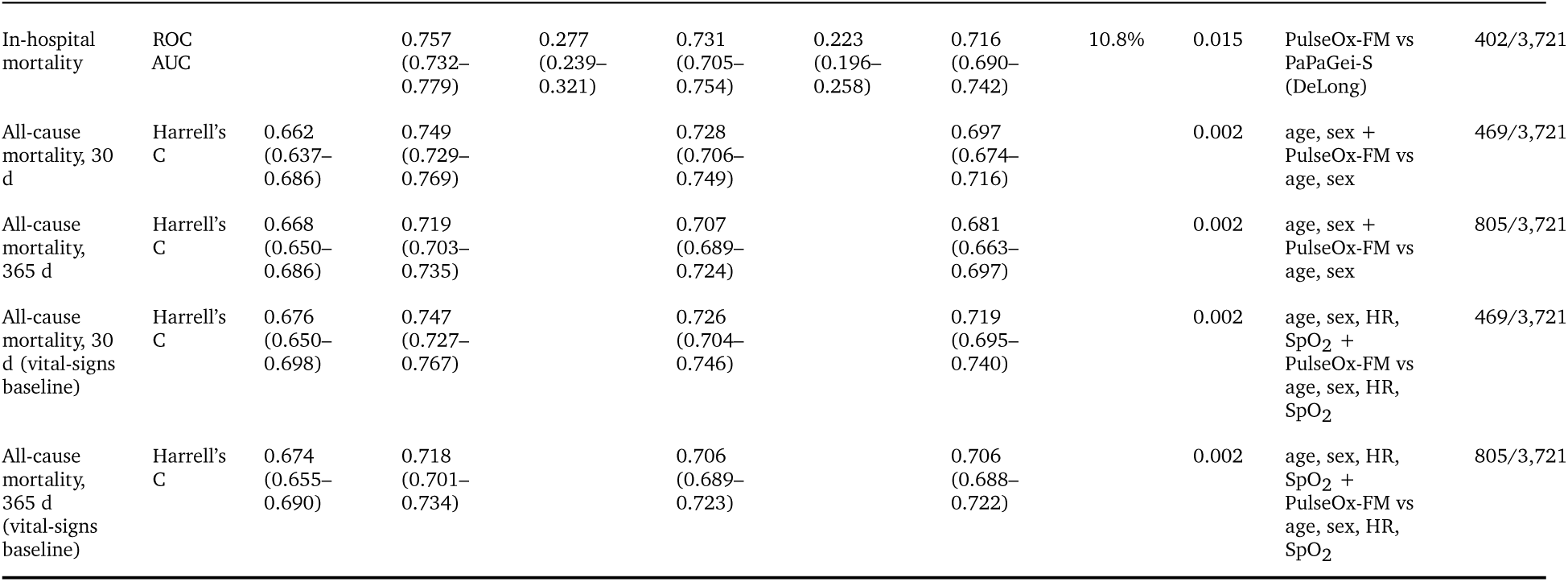
Numerical results for the prediction of health targets in HPP and VitalDB, and of mortality in MIMIC-III. Per-target performance for all tested HPP and VitalDB targets, including those not significantly improved (Figs 3a,b,d and 4a show the significant binary targets and every continuous target), computed as in those panels: one visit per participant, the earliest with both a labelled target and an embedding, with the recording as the unit of analysis and the participant as the cross-validation group. The primary metric is ROC AUC for binary targets and Pearson *r* for continuous targets, shown for demographics (age, sex and BMI) and for demographics plus pooled PulseOx-FM embeddings as the mean ± s.d. across three cross-validation seeds, with 95% bootstrap confidence intervals. For binary targets, the table also gives the area under the precision–recall curve (AUPRC) with the outcome prevalence, which is the AUPRC of a random classifier; for continuous targets, it gives R^2^. The *p* value is a two-sided paired bootstrap test of the improvement over demographics (2,000 participant-clustered resamples), and the *q* value is Benjamini–Hochberg-adjusted within each family (HPP continuous measures; HPP diagnoses and medications; VitalDB targets). n is the number of participants, or positives over participants for binary targets. The MIMIC-III block reports in-hospital mortality (ROC AUC and AUPRC, embeddings only) and 30-day and 365-day all-cause mortality (Harrell’s C, Cox models on age and sex, with PulseOx-FM embeddings added), each with a 95% bootstrap confidence interval. It also gives the PaPaGei-S comparator (pretrained on MIMIC-III), a randomly initialised encoder as a floor, and the comparison each *p* value tests. The two mortality blocks differ only in their comparator: the first adjusts for age and sex, the second (marked “vital-signs baseline”) for age, sex, mean heart rate and mean oxygen saturation, the vital signs a pulse oximeter itself reports. In every encoder column, the model is the stated covariates *plus* that encoder. BMI, body mass index; HR, heart rate; ROC AUC, receiver operating characteristic area under the curve; SpO_2_, peripheral oxygen saturation.

**Supplementary Table 5.** Numerical results for next-day target prediction from sleep embeddings versus sleep architecture. Per-target performance underlying Fig. 3f, listed by target domain (continuous glucose monitoring, CGM; dietary logging, Food; and wearable-derived activity, Wearables). The primary metric (Pearson *r*) is shown as mean ± standard deviation across cross-validation seeds for sleep-architecture features alone and for sleep architecture plus PulseOx-FM embeddings. The number of participants and nights, the combined-versus-architecture *p*-value and Benjamini–Hochberg FDR *q*-value, and the combined-model *p*-value and *q*-value are listed for each target. CGM, continuous glucose monitoring; CV, coefficient of variation; FDR, false discovery rate; GMI, glucose management indicator; PPGR, postprandial glucose response; SD, standard deviation; TAR, time above range; TBR, time below range; TIR, time in range.

| Domain | Target | Participants (n) | Nights (n) | Sleep architecture, Pearson $r$ | Sleep architecture + PulseOx-FM, Pearson $r$ | $P$, combined vs architecture | $q$, combined vs architecture | $P$, combined model | $q$, combined model |
| --- | --- | --- | --- | --- | --- | --- | --- | --- | --- |
| CGM | GMI | 6616 | 15892 | 0.027 ± 0.004 | 0.234 ± 0.003 | $1.7 \times 10^{-4}$ | $5.4 \times 10^{-4}$ | $2.4 \times 10^{-206}$ | $3.0 \times 10^{-205}$ |
| CGM | Mean glucose | 6616 | 15892 | 0.027 ± 0.004 | 0.234 ± 0.003 | $1.6 \times 10^{-4}$ | $5.4 \times 10^{-4}$ | $2.9 \times 10^{-206}$ | $3.0 \times 10^{-205}$ |
| CGM | PPGR 95th | 6616 | 15892 | 0.052 ± 0.003 | 0.147 ± 0.004 | 0.001 | 0.002 | $1.6 \times 10^{-79}$ | $6.8 \times 10^{-79}$ |
| CGM | Glucose SD | 6616 | 15892 | 0.058 ± 0.001 | 0.141 ± 0.006 | 0.002 | 0.003 | $6.9 \times 10^{-75}$ | $2.4 \times 10^{-74}$ |
| CGM | Glucose CV | 6616 | 15892 | 0.046 ± 0.001 | 0.107 ± 0.004 | 0.001 | 0.002 | $3.7 \times 10^{-41}$ | $6.0 \times 10^{-41}$ |

| Domain | Target | Participants<br>(n) | Nights<br>(n) | Sleep<br>architecture,<br>Pearson $r$ | Sleep architecture +<br>PulseOx-FM,<br>Pearson $r$ | $P$, combined<br>vs<br>architecture | $q$, combined<br>vs<br>architecture | $P$, combined<br>model | $q$, combined<br>model |
| --- | --- | --- | --- | --- | --- | --- | --- | --- | --- |
| CGM | TAR >180 | 6616 | 15892 | $0.024 \pm 0.003$ | $0.103 \pm 0.025$ | 0.021 | 0.023 | $1.5 \times 10^{-42}$ | $2.6 \times 10^{-42}$ |
| CGM | TIR 70–140 | 6616 | 15892 | $0.038 \pm 0.004$ | $0.087 \pm 0.006$ | 0.004 | 0.005 | $6.7 \times 10^{-30}$ | $8.8 \times 10^{-30}$ |
| CGM | TBR <70 | 6616 | 15892 | $-0.003 \pm 0.001$ | $0.047 \pm 0.003$ | $9.3 \times 10^{-4}$ | 0.002 | $1.2 \times 10^{-9}$ | $1.3 \times 10^{-9}$ |
| CGM | TIR 70–180 | 6616 | 15892 | $0.005 \pm 0.002$ | $0.043 \pm 0.003$ | 0.007 | 0.007 | $1.8 \times 10^{-8}$ | $1.9 \times 10^{-8}$ |
| Food | Energy | 3637 | 9968 | $0.035 \pm 0.004$ | $0.217 \pm 0.001$ | $9.7 \times 10^{-5}$ | $5.4 \times 10^{-4}$ | $1.3 \times 10^{-107}$ | $8.9 \times 10^{-107}$ |
| Food | Protein | 3637 | 9968 | $0.035 \pm 0.004$ | $0.206 \pm 0.001$ | $1.0 \times 10^{-4}$ | $5.4 \times 10^{-4}$ | $2.1 \times 10^{-95}$ | $1.1 \times 10^{-94}$ |
| Food | Fat | 3637 | 9968 | $-0.001 \pm 0.005$ | $0.176 \pm 0.002$ | $2.9 \times 10^{-4}$ | $7.5 \times 10^{-4}$ | $1.0 \times 10^{-70}$ | $3.0 \times 10^{-70}$ |
| Food | Fiber | 3637 | 9968 | $0.042 \pm 0.006$ | $0.162 \pm 0.003$ | $1.8 \times 10^{-4}$ | $5.4 \times 10^{-4}$ | $1.3 \times 10^{-60}$ | $2.6 \times 10^{-60}$ |
| Food | Weight | 3637 | 9968 | $0.031 \pm 0.003$ | $0.130 \pm 0.004$ | $1.5 \times 10^{-4}$ | $5.4 \times 10^{-4}$ | $3.4 \times 10^{-39}$ | $5.0 \times 10^{-39}$ |
| Food | Carbohydrate | 3637 | 9968 | $0.042 \pm 0.003$ | $0.122 \pm 0.0002$ | $1.2 \times 10^{-4}$ | $5.4 \times 10^{-4}$ | $4.4 \times 10^{-35}$ | $6.1 \times 10^{-35}$ |
| Food | Water | 3637 | 9968 | $0.032 \pm 0.002$ | $0.107 \pm 0.005$ | 0.001 | 0.002 | $1.3 \times 10^{-26}$ | $1.6 \times 10^{-26}$ |
| Food | Alcohol | 3637 | 9968 | $0.003 \pm 0.004$ | $0.100 \pm 0.003$ | 0.001 | 0.002 | $1.5 \times 10^{-23}$ | $1.8 \times 10^{-23}$ |
| Food | Caffeine | 3637 | 9968 | $-0.013 \pm 0.005$ | $-0.005 \pm 0.001$ | 0.041 | 0.041 | 0.606 | 0.606 |
| Wearables | Basal energy | 1117 | 3245 | $0.050 \pm 0.017$ | $0.312 \pm 0.010$ | 0.001 | 0.002 | $7.5 \times 10^{-69}$ | $2.0 \times 10^{-68}$ |
| Wearables | Active energy | 1208 | 3552 | $0.038 \pm 0.008$ | $0.249 \pm 0.010$ | $8.6 \times 10^{-4}$ | 0.002 | $1.5 \times 10^{-50}$ | $2.9 \times 10^{-50}$ |
| Wearables | Step count | 2803 | 9615 | $0.029 \pm 0.004$ | $0.149 \pm 0.003$ | 0.001 | 0.002 | $2.0 \times 10^{-66}$ | $4.7 \times 10^{-66}$ |

## Methods

### Description of cohort

The Human Phenotype Project (HPP) data presented in this paper was collected between January 2019 and May 2025, from a total of 11,311 participants, who were enrolled and underwent at least one home sleep test as part of the study. The study was approved by the Institutional Review Board (IRB) of the Weizmann Institute of Science (Reference number: 1719-1). The cohort in this study is healthy at the time of the recruitment (i.e. severe medical conditions and diseases were defined as exclusion criteria) and has follow-up visits every two years, which is planned for a total of 25 years. For full study design see ^22^. Of the 11,311 enrolled participants, who define the pretraining corpus and data split, 10,704 with a usable recording form the analysis cohort of the downstream evaluations, and 607 form the held-out test set (606 with a valid age prediction); per-target counts are given in the Supplementary Tables.

Sleep data were acquired using a home sleep apnea test (HSAT) device (WatchPAT 300; ZOLL Itamar), worn on the wrist and connected to a finger probe, capturing peripheral arterial tonometry (PAT), pulse oximetry (SpO_2_), heart rate, and actigraphy over the full night. In addition to sleep monitoring, participants underwent comprehensive multi-modal phenotyping including: blood tests (hematology, biochemistry), body composition assessment (dual-energy X-ray absorptiometry, DXA; anthropometrics), continuous glucose monitoring (CGM), dietary intake logging, wearable activity monitoring, fundus imaging, carotid and liver ultrasound, and health questionnaires capturing self-reported medical conditions and medication use. The raw red and near-infrared photoplethysmograms of the oximetry sensor are not part of the recordings exported by the device, which provide the derived SpO_2_ and pulse-rate series; the waveform channel used by the model is the infrared signal of the PAT finger probe. The sleep parameters used as prediction targets (AHI, RDI, ODI, sleep efficiency, total sleep and wake time, REM latency and sleep-stage proportions) are outputs of the device’s proprietary algorithm, validated against in-laboratory polysomnography ^20^, and are therefore reference labels of limited accuracy derived from the same signal as the embeddings.

### External Cohort

VitalDB ^23^ is a publicly available high-fidelity perioperative biosignal dataset comprising 6,388 surgical cases from Seoul National University Hospital, South Korea. Each case contains synchronous multi-parameter waveforms sampled at 500 Hz, including plethysmography (PPG/SpO_2_), ECG, and arterial pressure, along with vital-sign numerics recorded at 1–7 second intervals; on average 87 signal tracks per case are available. The dataset is IRB-approved (Seoul National University Hospital; NCT02914444) and released under CC BY-NC-SA 4.0. PulseOx-FM was applied to the plethysmography, heart rate and SpO_2_ channels of VitalDB recordings without any fine-tuning, providing an out-of-distribution test of model generalizability to a clinical (intraoperative) setting.

### Intensive-care cohort (MIMIC-III)

The MIMIC-III Waveform Database Matched Subset ^24^ links bedside-monitor waveform records of adult intensive-care patients at a tertiary academic centre in Boston, USA, to the MIMIC-III Clinical Database (v1.4). Records were linked to patients through the subject identifier; because MIMIC-III applies a single date shift per patient, intervals between waveform and clinical timestamps are preserved. We used the plethysmogram at its native 125 Hz, together with the heart-rate and SpO_2_ series of the accompanying numerics records (about 1 Hz), linearly interpolated onto the waveform grid. Non-overlapping 120-second windows were drawn from within contiguous recording segments; windows with more than 50% non-finite PPG samples or a flat signal were rejected, remaining missing samples were linearly interpolated, and up to 30 windows were taken per record. Each window was z-scored per channel and embedded with the frozen encoder, and up to 40 windows per patient, sampled evenly, were mean-pooled into a patient-level embedding. Patients with a plethysmographic channel, at least one usable window, clinical labels and an embedding from every encoder compared were eligible (n = 6,999). To ensure that no model input came from later in the stay or from a subsequent admission, we applied a 24-hour landmark: only waveform records beginning between 6 h before and 24 h after the first hospital admission contributed windows, and patients without such a record were excluded, leaving 3,721 patients (4,537 records; 132,711 windows). Age was taken at the first admission; because MIMIC-III date-shifts ages above 89 years, the 199 older patients were excluded from age prediction (n = 3,522) and entered the survival models with a training-fold median age and a missingness indicator. Outcomes were in-hospital death during the first admission (402 deaths) and all-cause death within 30 days (469) and 365 days (805) of the first admission, from the recorded date of death; patients without a recorded death were considered alive and censored at the horizon. BMI is not available in this cohort.

### Train, validation and test split

9,307,932 2-minutes segments, from 45,068 overnight recordings, collected during sleep monitoring of 11,311 participants in total, were randomly split by participants:

- Train: 6,995,558 2-minutes segments, from 33,877 overnight recordings, from 8,563 participants
- Validation: 1,741,098 2-minutes segments, from 8,405 overnight recordings from 2,141 participants
- Test: 571,276 2-minutes segments, from 2,786 overnight recordings from 607 participants

SSL pretraining was performed on recordings of the train set. Hyper-parameters optimization (batch size, learning rate, patch size, embedding dimension) for the pre-training was performed using a subset (10,000 segments) for each of the train and validation sets. Ablation experiments were performed on a subset of 1000 recordings from the train (for training) and validation (for evaluation) sets. The test set was entirely held out of the pretraining process including hyper-parameter tuning.

Reconstruction evaluations were evaluated on the held-out test set.

Downstream predictions were evaluated using 5 fold cross-validation, grouped by participants on either train and validation sets (42,282 recordings from 10,704 participants) or on the held-out test set (2,786 recordings from 607 participants).

### Preprocessing

The data was preprocessed using fixed-length segments at 125Hz sampling, that includes three synchronous channels: pulse-oximetry derived SpO_2_ (upsampled to 125Hz using temporal interpolation), heart rate (upsampled to 125hz, using temporal interpolation) and PAT infrared photoplethysmography (PPG, preprocessed and downsampled to 125Hz using the PyPPG pipeline ^27^). Input normalization used per-segment, per-channel z-scoring over time.

### Self-supervised learning architecture and pretraining

The model was trained to reconstruct masked portions of the 2-minute physiological segments, using a transformer-based masked-auto-encoder architecture ^50^adapted for multi-channel 1D input data (here physiological signals). The model’s encoder is based on 24 ViT ^51^ transformer blocks and decoder on 8 ViT transformer blocks, each block has 16 attention heads. The temporal ordering of patches along the time axis was encoded using fixed (non-learnable) sinusoidal positional embeddings ^52^, added to the patch token sequence in both the encoder and decoder, with the class token assigned a zero positional embedding. The optimization objective was a composite reconstruction Mean Squared Error (MSE) and Dynamic Time Warping (DTW) loss ^53^:

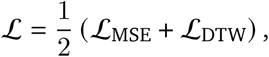

where 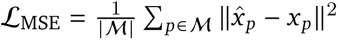 is the MSE over masked patches *M*, and *L*_DTW_ is the soft DTW distance between predicted and target masked-patch sequences. Both terms are applied per channel with channel-specific weights, selecting only the PPG/PAT loss channel.

Hyper-parameters: batch=128; learning rate=0.001, patch size=125 (1sec), embedding dimension=256, masking ratio= 0.5, input length=120sec.

Model optimization used AdamW ^54^ with weight decay 0.05, *β*_1_=0.9, *β*_2_=0.95, and a cosine annealing learning rate schedule with a 1-epoch linear warmup. The effect of masking ratio on pretraining quality was evaluated across both random masking reconstruction and forecasting tasks, demonstrating robust performance across a range of ratios and justifying the choice of 0.5 (Supplementary Figure 3).

Runtime - 4 weeks on 1 L40S NVIDIA GPUs

### Signal reconstruction evaluation

We benchmarked the PulseOx-FM model ability to reconstruct the PPG waveform, against a baseline model using classical signal processing and Fourier analysis. The baseline reconstructs each masked patch as a sinusoid fitted to the visible portions of the signal:

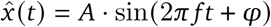

where *A* is the Root Mean Squared (RMS) amplitude of the visible patches, *f* is the normalized dominant frequency from the Fast Fourier Transform (FFT) of the visible signal (mean component excluded), and *φ* is the FFT phase at the dominant frequency.

PulseOx-FM demonstrated high fidelity in both forecasting and random masking tasks, achieving Pearson correlations of 0.99 ± 0.00 for forecasting (0s horizon) and 0.60 ± 0.02 for random masking (10% ratio), significantly outperforming the baseline (0.56 ± 0.06 and 0.15 ± 0.03, respectively) in capturing the repetitive yet complex structure of the PPG waveform (Extended Data Fig. 1 and Supp. Figure 1).

### Ablation experiments

Ablation experiments were performed on a subset of 1,000 recordings from the train and validation sets to characterize how key design choices affect downstream predictive accuracy. Age prediction performance was used as the primary readout, evaluated via 5-fold cross-validation with linear probing on frozen embeddings (Extended Data Fig. 2). We examined three axes: (i) pretraining duration (training epochs), (ii) input segment length (10, 30, 60, and 120 seconds), and (iii) masking ratio during pretraining (0.1–0.9). Ablation experiments revealed that predictive accuracy for global health markers scaled with input length, with 120-second segments providing significantly higher performance compared to shorter 10-second windows (Extended Data Fig. 2).

### Downstream predictions

To extract a subject-level embedding from overnight/surgical recordings, PulseOx-FM uses a “patch” approach: raw waveform segments are divided into fixed-length patches, encoded by the transformer into a sequence of tokens. A dedicated class token, prepended to the patch sequence and updated through the full depth of the transformer via self-attention, aggregates information across all patches into a single 256-dimensional latent vector representing the physiological state of the individual for that segment. Segment-level embeddings are then mean-pooled across the full overnight recording to produce a single subject-level representation (Extended Data Fig. 3). A prediction head, in our case *TabICLv2* python package ^55^, trained on these frozen embeddings was used to predict age or any other health target (current measured phenotypes, medication intake or disease) across all evaluations.

### Benchmark against PaPaGei-S

PaPaGei-S ^11^ was applied, with its released weights frozen, to the PPG channel of the same recordings: the HPP PAT waveform, the VitalDB plethysmogram resampled from 500 to 125 Hz, and the MIMIC-III plethysmogram. Following its published preprocessing, each 120-second segment was band-pass filtered (fourth-order Chebyshev type II, 0.5–12 Hz, zero-phase), smoothed with a 50-ms moving average, z-scored and divided into twelve 10-second windows. The 512-dimensional window embeddings were averaged per segment and then per recording or patient, as for PulseOx-FM. Both encoders were evaluated on the same recording intersection, with the same participant-grouped folds and the same prediction heads, so that the comparison measures the difference between models rather than between pipelines. PaPaGei was pretrained on public datasets that include VitalDB and MIMIC-III, which are therefore in-distribution for PaPaGei-S but out-of-distribution for PulseOx-FM. In MIMIC-III, an encoder with the PulseOx-FM architecture and random weights served as a floor.

### Evaluation metrics and statistical analysis

To evaluate the dataset, we implemented a subject-level cross-validation (CV) framework. Data splitting was performed strictly at the subject level to prevent data leakage between visits. We employed a 5-fold outer loop for performance estimation, repeated across 3 distinct random seeds (generating n=15 independent train-test splits) to produce stable mean estimates and confidence intervals.

The predictive power for each model was reported as the mean metric (Pearson correlation coefficient (r) and coefficient of determination (R^2^) for continuous regression targets; Area Under the Receiver Operating Characteristic Curve (AUC-ROC) for binary classification) across the seeds of predicted folds. Where applicable, Bland–Altman agreement (age prediction) and the area under the precision– recall curve (AUPRC; binary targets) were also computed.

To rigorously validate associations, we compared the predictive performance of the full model, i.e. embeddings concatenated with demographic covariates (age, sex and BMI), against the baseline model of covariates only.

An association was considered statistically significant only if the full model’s performance distribution was significantly higher than that of the baseline after Benjamini–Hochberg FDR correction (FDR q < 0.10).

Improvements over demographics were tested bya two-sided paired bootstrap over 2,000 participant-clustered resamples, with 95% percentile confidence intervals, and Benjamini–Hochberg correction was applied within three families (HPP continuous measures; HPP diagnoses and medications; VitalDB targets). Effect sizes are reported with 95% confidence intervals throughout, per-target results carry Benjamini–Hochberg *q* values, and Supp. Table 3 reports the mean and standard deviation across cross-validation seeds.

### Agreement and the PulseOx-age gap

Agreement between predicted and chronological age was assessed by Bland–Altman analysis ^56^ on the seed-averaged out-of-fold predictions, with the difference between predicted and chronological age plotted against chronological age, the error-free reference, rather than against the mean of the two ^57^. Bias, 95% limits of agreement (bias ± 1.96 s.d.) and the proportion of participants within them are reported for every model and cohort (Supp. Table 3). For association analyses, the dependence of the gap on chronological age was removed with the slope-and-intercept correction of de Lange and Cole ^58^: predicted age was regressed on chronological age (predicted = *α* + *β* × age) and each prediction mapped to (predicted - *α*)/*β*, with *α* and *β* estimated on the training folds only and applied to the held-out fold (five participant-level folds, three seeds, corrected values averaged across seeds). The bias-corrected gap, the corrected prediction minus chronological age, was standardised and related at baseline visits to systolic and diastolic blood pressure, triglycerides, HDL cholesterol, fasting glucose, waist circumference and carotid intima-media thickness (linear regression on standardised outcomes) and to metabolic syndrome (logistic regression), adjusted for age, age^2^, sex and BMI, with Wald 95% confidence intervals and Benjamini–Hochberg correction across the eight endpoints. Metabolic syndrome was defined by the harmonised criteria ^37^ as at least three of: waist circumference ≥ 102 cm in men or ≥ 88 cm in women; triglycerides ≥ 150 mg/dL; HDL cholesterol < 40 mg/dL in men or < 50 mg/dL in women; systolic pressure ≥ 130 mmHg, diastolic pressure ≥ 85 mmHg or antihypertensive treatment; and fasting glucose ≥ 100 mg/dL or antidiabetic treatment. To compare accelerated with decelerated agers, participants were split by quartile of the age-gap residual (the residual of predicted on chronological age), and the top and bottom quartiles, age-matched by construction, were compared on every phenotype with at least 50 participants per group and 10 positive cases: continuous measures by Cohen’s *d* and two-sided Mann–Whitney *U* test, and binary targets by log_2_ odds ratio and two-sided Fisher exact test, with 95% confidence intervals from 2,000 bootstrap resamples and Benjamini–Hochberg correction within each task family (*q* < 0.05).

### Subgroup analyses

In HPP, ancestry was derived from the questionnaire-reported country of birth of each participant’s four grandparents. A participant was assigned to a country when all grandparents with an informative country of birth were born there (grandparents born in Israel were considered uninformative), and countries were grouped into six ancestry groups (Ashkenazi, Sephardi, Middle Eastern, North African, Yemenite and other). Participants whose grandparents came from different groups were labelled mixed. This coded 8,678 of 9,443 participants (91.9%). Within each group, Pearson *r* and mean absolute error were computed from the out-of-fold age predictions of PulseOx-FM and of PaPaGei-S, fitted under the identical protocol, with 95% participant-clustered bootstrap confidence intervals. Systematic bias by ancestry was tested by regressing the signed prediction error on ancestry group, adjusted for age and age^2^, with a participant-clustered Wald test. In MIMIC-III, the C-index of the embeddings-only Cox models at 30 and 365 days was computed within self-reported ethnicity (five groups) and ten-year age bands, with 95% confidence intervals from 500 patient-level bootstrap resamples; subgroups with fewer than five deaths were omitted. Heterogeneity was tested by a likelihood-ratio test for the interaction between the predicted risk score and the subgroup in a Cox model.

### Survival and decision-curve analysis

Survival in MIMIC-III was modelled with Cox proportional-hazards models (lifelines) on covariate sets representing the information available at the bedside: age and sex; age and sex plus the mean heart rate and mean oxygen saturation pooled over the same 120-s windows the encoder read; pooled embeddings alone; and each covariate set plus embeddings. Embedding models were ridge-penalised, with the penalty selected by threefold inner cross-validation within each training fold to maximise concordance. Models were fitted within the participant-grouped five-fold cross-validation repeated over three seeds, and all reported metrics use out-of-fold predictions. Discrimination was measured by Harrell’s C-index, with 95% confidence intervals from 1,000 patient-level bootstrap resamples, and differences between models were tested by a two-sided paired bootstrap on the same resamples. For survival curves, patients were ranked by held-out predicted risk and divided into quartiles; Kaplan–Meier curves with Greenwood 95% confidence intervals were compared between each model’s lowest and highest quartile by log-rank test, with Benjamini–Hochberg adjustment across models and horizons. In-hospital death was predicted by the TabICLv2 classifier from the embeddings; ROC AUC and AUPRC are reported with 95% confidence intervals from 2,000 bootstrap resamples, and encoders were compared by paired bootstrap and DeLong’s test. Cumulative incidence of in-hospital death by quartile of predicted risk was estimated with the Aalen–Johansen estimator, treating discharge alive as a competing event rather than as censoring ^59^, with 95% bands from 1,000 patient-level bootstrap resamples and a log-rank test for trend. For decision-curve analysis ^60^, out-of-fold linear predictors were converted to absolute risks at each horizon through a baseline cumulative hazard cross-fitted over the same folds, set so that expected events equalled observed events in the training folds. Net benefit, NB = TP/*n* - (FP/*n*) × *p*_t_/(1 - *p*_t_), was computed over risk thresholds *p*_t_ from 1% to 60% and compared with treating all and no patients, with 95% bands from 2,000 patient-level bootstrap resamples. Because the average predicted risk is fixed to the observed event rate, these curves do not assess calibration in the large.

### Two-year incident hypertension

Incident hypertension was assessed in participants with a baseline and a two-year follow-up visit who were normotensive at baseline (systolic < 130 mmHg and diastolic < 85 mmHg), untreated and without a reported hypertension diagnosis. Incident hypertension was defined as systolic ≥ 140 mmHg or diastolic ≥ 90 mmHg at the follow-up visit, antihypertensive treatment, or a reported diagnosis with onset within two years of baseline (n = 3,714; 179 cases). Predictions came from the TabICLv2 head in participant-level five-fold cross-validation repeated over three seeds. For Fig. 3c, age, sex and BMI were compared with and without the embeddings, using the mean over seeds of the ROC AUC, 95% participant bootstrap confidence intervals and a two-sided paired bootstrap test (2,000 resamples), outside the FDR families. On the same participants, the embeddings were also added to a standard-of-care comparator of age, sex, BMI and baseline systolic and diastolic blood pressure; AUROCs were compared by DeLong’s test, and decision curves were evaluated over risk thresholds from 5% to 25%.

### Temporal stability

Temporal stability (Extended Data Fig. 4) was assessed by predicting age from embeddings of a single time bin and from embeddings pooled cumulatively from the start of monitoring: two-minute segments up to 8 h in HPP, 4 h in VitalDB and 1 h in MIMIC-III, in patients with a record in the first day.

## Data availability

Data in this paper is part of the Human Phenotype Project (HPP) and is accessible to researchers from universities and other research institutions at: https://humanphenotypeproject.org/data-access. The VitalDB dataset used for external validation is publicly available (CC BY-NC-SA 4.0) at https://vitaldb.net. The MIMIC-III Clinical Database and Waveform Database Matched Subset are available to credentialed users through PhysioNet (https://physionet.org).

## Code availability

All code, including preprocessing, pretraining and model definition, is available at https://github.com/SarahKohn/PulseOx-FM. Upon publication, the pretrained PulseOx-FM weights will be made available to researchers through the Human Phenotype Project data-access process (https://humanphenotypeproject.org/data-access).

## Acknowledgments

We thank the members of the Segal group for fruitful discussions. S.K is a fellow of the Ariane de Rothschild Women Doctoral Excellence Program. E.S. is supported by the Crown Human Genome Center; the Larson Charitable Foundation New Scientist Fund; the Else Kröner Fresenius Foundation; the White Rose International Foundation; the Ben B. and Joyce E. Eisenberg Foundation; the Nissenbaum Family; Marcos Pinheiro de Andrade and Vanessa Buchheim; Lady Michelle Michels; Aliza Moussaieff; and grants funded by the Minerva Foundation, with funding from the Federal German Ministry for Education and Research and by the European Research Council and the Israel Science Foundation. Funded by the European Union Project 101095540 IMMEDIATE. Views and opinions expressed are, however, those of the author(s) only and do not necessarily reflect those of the European Union. Neither the European Union nor the granting authority can be held responsible for them. The funders had no role in study design, data collection and analysis, decision to publish or preparation of the manuscript.

## Author Contributions

S.K. conceived the project, designed the model and conducted all analyses, interpreted the results and wrote the manuscript. G.L. and A.D. reviewed the analyses, results and manuscript. S.S. interpreted the results and reviewed the manuscript. A.G. performed clinical data preprocessing. A.G., G.S., G.W. and A.W. contributed to the analysis code. A.W. developed protocols and oversaw sample collection and processing. E.S. and H.R conceived and directed the project, designed the analyses, reviewed the results and the manuscript.

## References

1. Park, J., Seok, H. S., Kim, S.-S. & Shin, H. Photoplethysmogram analysis and applications: an integrative review. Front. Physiol. 12, 808451 (2021).

2. [2401.12783] A Scoping Review of Deep Learning Methods for Photoplethysmography Data. https://arxiv.org/abs/2401.12783.

3. Singh, S., Khan, S. Z., Singh, D., Verma, S. & Talwar, A. The uses of overnight pulse oximetry. Lung India 37, 151–157 (2020).

4. Elgendi, M. On the analysis of fingertip photoplethysmogram signals. Curr. Cardiol. Rev. 8, 14–25 (2012).

5. Li, R. et al. Large-Scale Validation of a Dual Cross-Attention Network for Automated Sleep Staging Using Wearable Photoplethysmography Signals. Diagnostics (Basel) 16, (2026).

6. Bommasani, R., et al. On the Opportunities and Risks of Foundation Models. arXiv (2021) doi:10.48550/arxiv.2108.07258.

7. Zhou, Y. et al. A foundation model for generalizable disease detection from retinal images. Nature 622, 156–163 (2023).

8. Chen, R. J. et al. Towards a general-purpose foundation model for computational pathology. Nat. Med. 30, 850–862 (2024).

9. Lutsker, G. et al. A foundation model for continuous glucose monitoring data. Nature 650, 978–986 (2026).

10. Abbaspourazad, S., et al. Large-scale Training of Foundation Models for Wearable Biosignals. arXiv (2023) doi:10.48550/arxiv.2312.05409.

11. Pillai, A., Spathis, D., Kawsar, F. & Malekzadeh, M. PaPaGei: Open Foundation Models for Optical Physiological Signals. arXiv (2024) doi:10.48550/arxiv.2410.20542.

12. Saha, M. et al. Pulse-PPG: An Open-Source Field-Trained PPG Foundation Model for Wearable Applications across Lab and Field Settings. Proc. ACM Interact. Mob. Wearable Ubiquitous Technol. 9, 1–35 (2025).

13. Chen, Z. et al. GPT-PPG: a GPT-based foundation model for photoplethysmography signals. Physiol. Meas. 46, (2025).

14. [2511.01747] AnyPPG: An ECG-Guided PPG Foundation Model Trained on Over 100,000 Hours of Recordings for Holistic Health Profiling. https://arxiv.org/abs/2511.01747.

15. [2509.19215] PPG-Distill: Efficient Photoplethysmography Signals Analysis via Foundation Model Distillation. https://arxiv.org/abs/2509.19215.

16. Miller, A. C. et al. A wearable-based aging clock associates with disease and behavior. Nat. Commun. 16, 9264 (2025).

17. Nie, G., Zhao, Q., Tang, G., Li, Y. & Hong, S. Artificial intelligence-derived photoplethysmography age as a digital biomarker for cardiovascular health. Commun Med (London) 5, 481 (2025).

18. Thapa, R. et al. A Multimodal Sleep Foundation Model Developed with 500K Hours of Sleep Recordings for Disease Predictions. medRxiv (2025) doi:10.1101/2025.02.04.25321675.

19. Kohn, S. et al. Phenome-wide associations of sleep characteristics in the Human Phenotype Project. Nat. Med. 31, 1026–1037 (2025).

20. Körkuyu, E. et al. The efficacy of Watch PAT in obstructive sleep apnea syndrome diagnosis. Eur Arch Otorhinolaryngol 272, 111–116 (2015).

21. Attia, S. et al. Clinical Validation of Artificial Intelligence Algorithms for the Diagnosis of Adult Obstructive Sleep Apnea and Sleep Staging From Oximetry and Photoplethysmography-SleepAI. J. Sleep Res. 35, e70093 (2026).

22. Reicher, L. et al. Deep phenotyping of health-disease continuum in the Human Phenotype Project. Nat. Med. 31, 3191–3203 (2025).

23. Lee, H.-C. et al. VitalDB, a high-fidelity multi-parameter vital signs database in surgical patients. Sci. Data 9, 279 (2022).

24. Johnson, A. E. W. et al. MIMIC-III, a freely accessible critical care database. Sci. Data 3, 160035 (2016). doi:10.1038/sdata.2016.35

25. Hellqvist, H. et al. Overnight stiffness index from finger photoplethysmography in relation to markers of cardiovascular risk and vascular ageing. Heart Vessels 40, 895–904 (2025).

26. Charlton, P. H. et al. Assessing hemodynamics from the photoplethysmogram to gain insights into vascular age: a review from VascAgeNet. Am. J. Physiol. Heart Circ. Physiol. 322, H493–H522 (2022).

27. Goda, M. Á., Charlton, P. H. & Behar, J. A. pyPPG: a Python toolbox for comprehensive photoplethysmography signal analysis. Physiol. Meas. 45, (2024).

28. McInnes, L., Healy, J. & Melville, J. UMAP: Uniform Manifold Approximation and Projection for Dimension Reduction. arXiv (2018) doi:10.48550/arxiv.1802.03426.

29. Ujma, P. P. & Bódizs, R. Correlates of sleep variability in a mobile EEG-based volunteer study. Sci. Rep. 14, 26012 (2024).

30. Messman, B. A. et al. How much does sleep vary from night-to-night? A quantitative summary of intraindividual variability in sleep by age, gender, and racial/ethnic identity across eight-pooled datasets. J. Sleep Res. 31, e13680 (2022).

31. Doherty, C., Baldwin, M., Lambe, R., Burke, D. & Altini, M. Readiness, recovery, and strain: an evaluation of composite health scores in consumer wearables. Transl. Exerc. Biomed. (2025) doi:10.1515/teb-2025-0001.

32. Thapa, R. et al. A multimodal sleep foundation model for disease prediction. Nat. Med. 32, 752–762 (2026).

33. Vlachopoulos, C., O’Rourke, M. & Nichols, W. W. Mcdonald’s blood flow in arteries: theoretical, experimental and clinical principles. (CRC Press, 2011). 10.1201/b13568.

34. Millasseau, S. C. et al. Noninvasive assessment of the digital volume pulse. Comparison with the peripheral pressure pulse. Hypertension 36, 952–956 (2000).

35. Levine, M. E. et al. An epigenetic biomarker of aging for lifespan and healthspan. Aging (Albany NY) 10, 573–591 (2018).

36. Belsky, D. W. et al. Quantification of biological aging in young adults. Proc Natl Acad Sci USA 112, E4104–10 (2015).

37. Alberti, K. G. M. M. et al. Harmonizing the metabolic syndrome: a joint interim statement of the International Diabetes Federation Task Force on Epidemiology and Prevention; National Heart, Lung, and Blood Institute; American Heart Association; World Heart Federation; International Atherosclerosis Society; and International Association for the Study of Obesity. Circulation 120, 1640–1645 (2009).

38. Drager, L. F., Togeiro, S. M., Polotsky, V. Y. & Lorenzi-Filho, G. Obstructive sleep apnea: a cardiometabolic risk in obesity and the metabolic syndrome. J. Am. Coll. Cardiol. 62, 569–576 (2013).

39. Sipilä, K. et al. Metabolic syndrome and carotid intima media thickness in the Health 2000 Survey. Atherosclerosis 204, 276–281 (2009).

40. Targher, G., Valenti, L. & Byrne, C. D. Metabolic Dysfunction-Associated Steatotic Liver Disease. N. Engl. J. Med. 393, 683–698 (2025).

41. Chang, Y.-W., Hsiu, H., Yang, S.-H., Fang, W.-H. & Tsai, H.-C. Characteristics of beat-to-beat photoplethysmography waveform indexes in subjects with metabolic syndrome. Microvasc. Res. 106, 80–87 (2016).

42. Somers, V. K. et al. Sleep apnea and cardiovascular disease: an american heart association/american college of cardiology foundation scientific statement from the american heart association council for high blood pressure research professional education committee, council on clinical cardiology, stroke council, and council on cardiovascular nursing in collaboration with the national heart, lung, and blood institute national center on sleep disorders research (national institutes of health). Circulation 118, 1080–1111 (2008).

43. Pencina, M. J., D’Agostino, R. B., Larson, M. G., Massaro, J. M. & Vasan, R. S. Predicting the 30-year risk of cardiovascular disease: the framingham heart study. Circulation 119, 3078–3084 (2009).

44. Fiani, D., Campbell, H., Solmi, M., Fiedorowicz, J. G. & Calarge, C. A. Impact of antidepressant use on the autonomic nervous system: A meta-analysis and systematic review. Eur. Neuropsychopharmacol. 71, 75–95 (2023).

45. Carnethon, M. R. et al. Risk factors for progression to incident hyperinsulinemia: the Atherosclerosis Risk in Communities Study, 1987-1998. Am. J. Epidemiol. 158, 1058–1067 (2003).

46. Keller, M. D., Harrison-Smith, B., Patil, C. & Arefin, M. S. Skin colour affects the accuracy of medical oxygen sensors. Nature 610, 449–451 (2022). doi:10.1038/d41586-022-03161-1

47. Mancia, G. et al. 2023 ESH Guidelines for the management of arterial hypertension The Task Force for the management of arterial hypertension of the European Society of Hypertension: Endorsed by the International Society of Hypertension (ISH) and the European Renal Association (ERA). J. Hypertens. 41, 1874–2071 (2023).

48. Thase, M. E. Effects of venlafaxine on blood pressure: a meta-analysis of original data from 3744 depressed patients. J. Clin. Psychiatry 59, 502–508 (1998).

## Methods References

49. Paperclip. https://paperclip.gxl.ai/.

50. He, K., et al. Masked Autoencoders Are Scalable Vision Learners. arXiv (2021) doi:10.48550/arxiv.2111.06377.

51. Dosovitskiy, A., et al. An Image is Worth 16×16 Words: Transformers for Image Recognition at Scale. arXiv (2020) doi:10.48550/arxiv.2010.11929.

52. Vaswani, A., et al. Attention is all you need. arXiv (2017) doi:10.48550/arxiv.1706.03762.

53. Cuturi, M. & Blondel, M. Soft-DTW: a Differentiable Loss Function for Time-Series. arXiv (2017) doi:10.48550/arxiv.1703.01541.

54. Loshchilov, I. & Hutter, F. Decoupled Weight Decay Regularization. arXiv (2017) doi:10.48550/arxiv.1711.05101.

55. [2602.11139] TabICLv2: A better, faster, scalable, and open tabular foundation model. https://arxiv.org/abs/2602.11139.

56. Bland, J. M. & Altman, D. G. Statistical methods for assessing agreement between two methods of clinical measurement. Lancet 327, 307–310 (1986). doi:10.1016/S0140-6736(86)90837-8

57. Krouwer, J. S. Why Bland–Altman plots should use X, not (Y+X)/2 when X is a reference method. Stat. Med. 27, 778–780 (2008). doi:10.1002/sim.3086

58. de Lange, A.-M. G. & Cole, J. H. Commentary: Correction procedures in brain-age prediction. NeuroImage Clin. 26, 102229 (2020). doi:10.1016/j.nicl.2020.102229

59. Austin, P. C., Lee, D. S. & Fine, J. P. Introduction to the analysis of survival data in the presence of competing risks. Circulation 133, 601–609 (2016). doi:10.1161/CIRCULATIONAHA.115.017719

60. Vickers, A. J. & Elkin, E. B. Decision curve analysis: a novel method for evaluating prediction models. Med. Decis. Making 26, 565–574 (2006). doi:10.1177/0272989X06295361

